# miR-21 blocks obesity in mice: a potential therapy for humans

**DOI:** 10.1101/2020.10.27.20219915

**Authors:** Said Lhamyani, Adriana-Mariel Gentile, Rosa M. Giráldez-Pérez, Mónica Feijóo-Cuaresma, Silvana Yanina Romero-Zerbo, Mercedes Clemente-Postigo, Hatem Zayed, Wilfredo Oliva Olivera, Francisco Javier Bermúdez-Silva, Julián Salas, Carlos López Gómez, Nabil Hajji, Gabriel Olveira Fuster, Francisco J. Tinahones, Rajaa El Bekay

## Abstract

microRNAs are promising drug targets in obesity and metabolic disorders. miR-21 expression is upregulated in obese white adipose tissue (WAT); however, its physiological role in WAT has not been fully explored. We aimed to dissect the underlying molecular mechanisms of miR-21 in treating obesity, diabetes, and insulin resistance. We demonstrated, in human and mice, that elevated miR-21 expression is associated with metabolically healthy obesity. miR-21 mimic affected the expression of genes associated with adipogenesis, thermogenesis, and browning in 3T3-L1 adipocytes. In addition, it blocked high fat diet-induced weight gain in obese mice, without modifying food intake or physical activity. This was associated with metabolic enhancements, WAT browning and thermogenic programming, and brown AT induction through VEGF-A, p53, and TGFβ1 signaling pathways. Our findings add a novel role of miR-21 in the regulation of obesity and a potential therapy for both obesity and T2D without altering caloric intake and physical activities.

## Introduction

Obesity is slowly becoming a global health epidemic. It is commonly associated with different diseases, including diabetes, heart disease, stroke, and cancer, compromising the quality of life and putting an enormous economic burden on society (Hruby & Hu, 2015). However, there are no efficient therapies to date for the treatment of obesity. Recently, microRNAs (miRNAs) have arisen as potential therapeutic targets due to their regulatory role in many biological processes, including transcriptional regulation of metabolism (Zhong *et al*, 2018).

It is well established that white adipose tissue (WAT) dysfunction leads to the development of obesity-related metabolic disturbances (Rodríguez *et al*, 2015; Chouchani & Kajimura, 2019). Given the existence of metabolically healthy obese subjects and diabetic lean individuals, it has been suggested that when the expansion capacity of adipose tissue (AT) required to store excess energy is exceeded, metabolic disorders occur (Hajer *et al*, 2008). In this case, dysfunctional WAT results in the accumulation of fat in non-fatty organs, such as muscles, the pancreas, liver, and the heart, leading to lipotoxicity and impaired insulin signaling (Friesen & Cowan, 2019; Chouchani & Kajimura, 2019).

In addition to WAT, brown (BAT) and beige AT, rich in mitochondria, are involved in weight management and metabolic homeostasis due to their high energy-utilizing capacity through thermogenesis (Harms & Seale, 2013; Villarroya & Vidal-Puig, 2013). Beige thermogenic adipocytes arise in WAT depots after continuous exposure to cold, β3-adrenergic stimulation or by genetic manipulation of certain specific pathways. This process is known as browning (Reitman, 2017), which can be induced pharmacologically by activation of the β3-adrenergic receptor and thiazolidinediones (Reitman, 2017). Interestingly, rodent models have shown that WAT browning was able to promote metabolic improvement and resistance to diet-induced obesity (Seale *et al*, 2011).

Within this context, interest has increased in the role of miRNAs in the development of fat cells and obesity (Zhong *et al*, 2018). In particular, many miRNAs have been defined as regulators of the differentiation and function of beige AT and BAT (Karbiener & Scheideler, 2014). Understanding the role of miRNAs in the thermogenic activation of BAT and the browning of WAT can provide new therapeutic targets against obesity and associated metabolic diseases. To take advantage of the full potential of miRNA-based therapies, selecting most suitable miRNA is required, which makes the performing of comprehensive studies on candidate miRNA function necessary.

miR-21 was found to be frequently upregulated in many chronic diseases, such as obesity(Sekar *et al*, 2014; Keller *et al*, 2011). miR-21 is upregulated in epididymal WAT from obese mice compared to normoweights (NW) (Keller *et al*, 2011), and in type 2 diabetic obese compared to non-diabetic obese subjects (Guglielmi *et al*, 2017). miR-21 enhances adipogenic differentiation through the modulation of transforming growth factor (TGF-β) signaling (Jeong Kim *et al*, 2009; Lee *et al*, 2011), and plays a pivotal role in angiogenesis through the regulation of vascular endothelial growth factor A (VEGF-A), which is in turn known to be a regulator of thermogenesis (Richart *et al*, 2014). However, the precise mechanisms underlying its relationship with obesity and type 2 diabetes or insulin resistance have not yet been described. Therefore, based on previous bioinformatics analysis, we aimed to analyze the effect of miR-21 on the principal processes involved in the regulation of WAT function and expansion. In this regard, through *in vivo* and *in vitro* experiments in which miR-21 effects are mimicked, we aimed to characterize the molecular mechanisms underlying these effects and the real connection between the increase of this miR-21, adipose tissue functionality, obesity, type 2 diabetes and insulin resistance.

## Results

### In-silico analysis of miR-21 potential targets

miR-21 has been suggested to be potentially related to obesity and to be involved in adipogenesis regulation (Keller *et al*, 2011). Therefore, it is important to investigate the real role of this miRNA in AT functionality regulation. It is well known that several factors such as angiogenesis, adipogenesis and apoptosis are related to adipose tissue functionality and then to obesity and associated metabolic alterations (Tinahones *et al*, 2013, 2012). We investigated the role of miR-21 in obesity using different bioinformatics tools as mentioned in the Methods section and 1505 validated target genes for miR-21, mainly involved in angiogenesis, VEGF signaling, apoptosis, adipogenesis and brown adipocyte differentiation, were identified (**Table S1 and Table S2**).

### The effect of miR-21 mimic on the expression levels of genes involved in angiogenesis, apoptosis, thermogenic and browning processes in 3T3-L1 differentiated adipocytes

To biologically validate the *in-silico* predicted miR-21 target genes, we investigated the effects of miR-21 mimic administration in 3T3-L1 differentiated adipocytes. *In vitro* treatment of 3T3-L1 cells with miR-21 mimic was validated by the significant increase of miR-21 levels in treated cells compared to controls (**Fig. 1A**). miR-21 mimic led to a significant increase in angiogenic genes, mainly *Vegf-A, Vegf-B* and *Vegf-C*, and decreased *Ang2, Ang4* and the anti-angiogenic *Timp-3* genes compared to control cells (**Fig 1B**). Moreover, miR-21 mimic led to the significant increase of both the anti-apoptotic *Bcl-2* and the pro-apoptotic *Casp-3* and *Bid*, compared to controls (**Fig.1C**). However, both *Ppar-g* and *Cebp-a* gene expressions were significantly decreased with miR-21 mimic treatment compared to control (**Fig. 1D**), while genes involved in BAT differentiation, thermogenesis, and browning, mainly *Ucp1, Fgf-21, Pgc-1a* and *Tmem26*, showed an increase with mimic treatment compared to controls (**Fig. 1E**).

**Figure 1.**
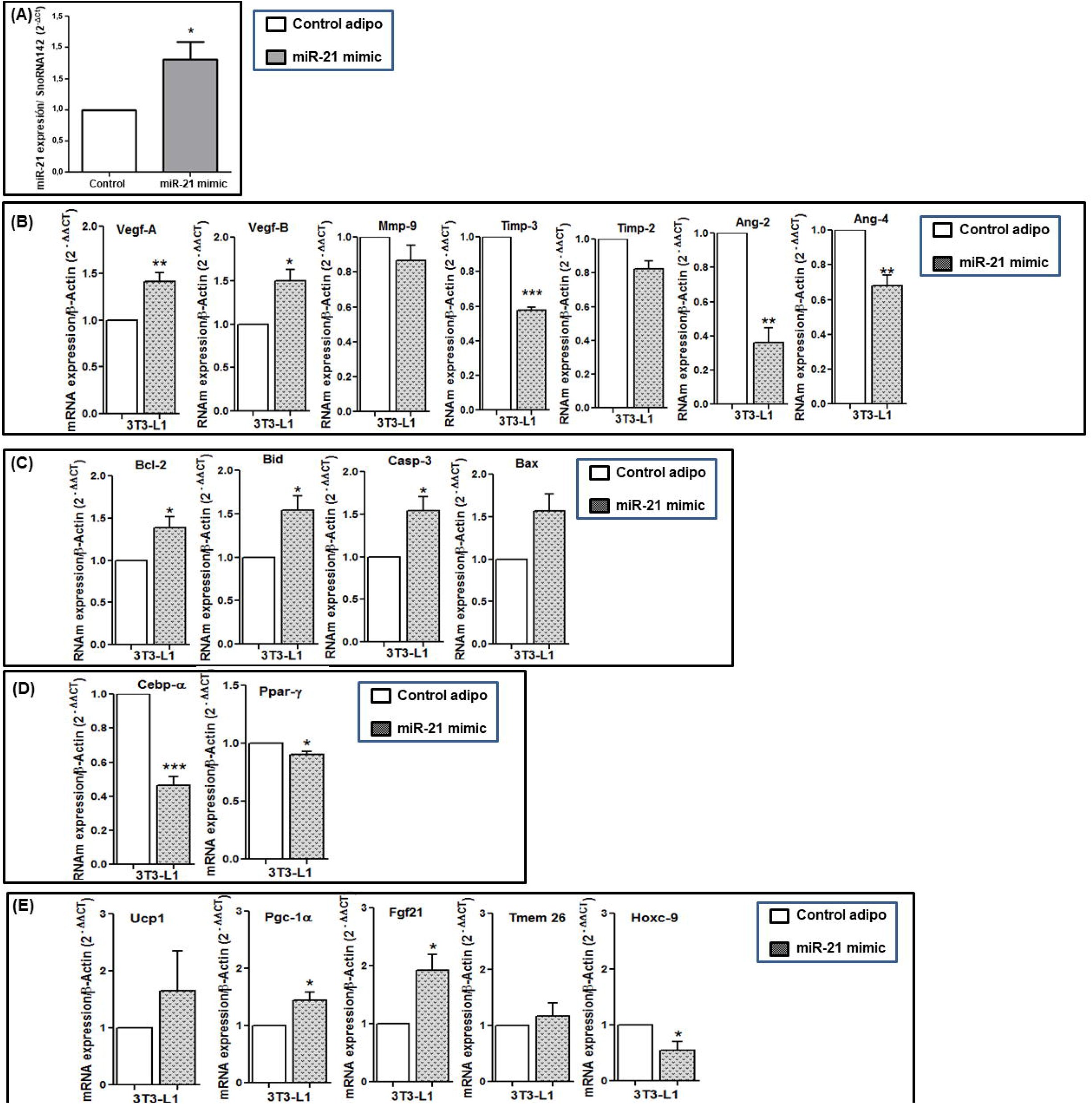
*In vitro* effect of miR-21 mimic on the mRNA expression of angiogenic, apoptotic, adipogenic, browning and thermogenesis markers in 3T3-L1 adipocytes. Differentiated 3T3-L1 cells were treated with miR-21 mimic 5 nM or control mimic (n=6 per group) for 48 hours. **(A)** miR-21 levels measured by real-time qPCR using SnoRNA142 as reference miRNA (2^ΔΔct^). Data are expressed as the mean ± SEM. *P<0.05 miR-21 mimic versus control according to the Student’s t-test. The expression levels of miR-21 were also measured in cell cultures in the presence of control mimic and transfect and no differences were observed compared to those obtained in control adipo (data not shown). mRNA levels of angiogenic **(B)**, apoptotic **(C)**, adipogenic **(D)** and browning and thermogenesis **(E)** genes were measured by real-time qPCR using β-ctin as a reference gene (2^-ΔΔct^). *p□0.05 and **p□0.01 versus control mimic. Data are expressed as the mean ± SEM. Student’s t-test was used for statistical analysis.

**Figure 2.**
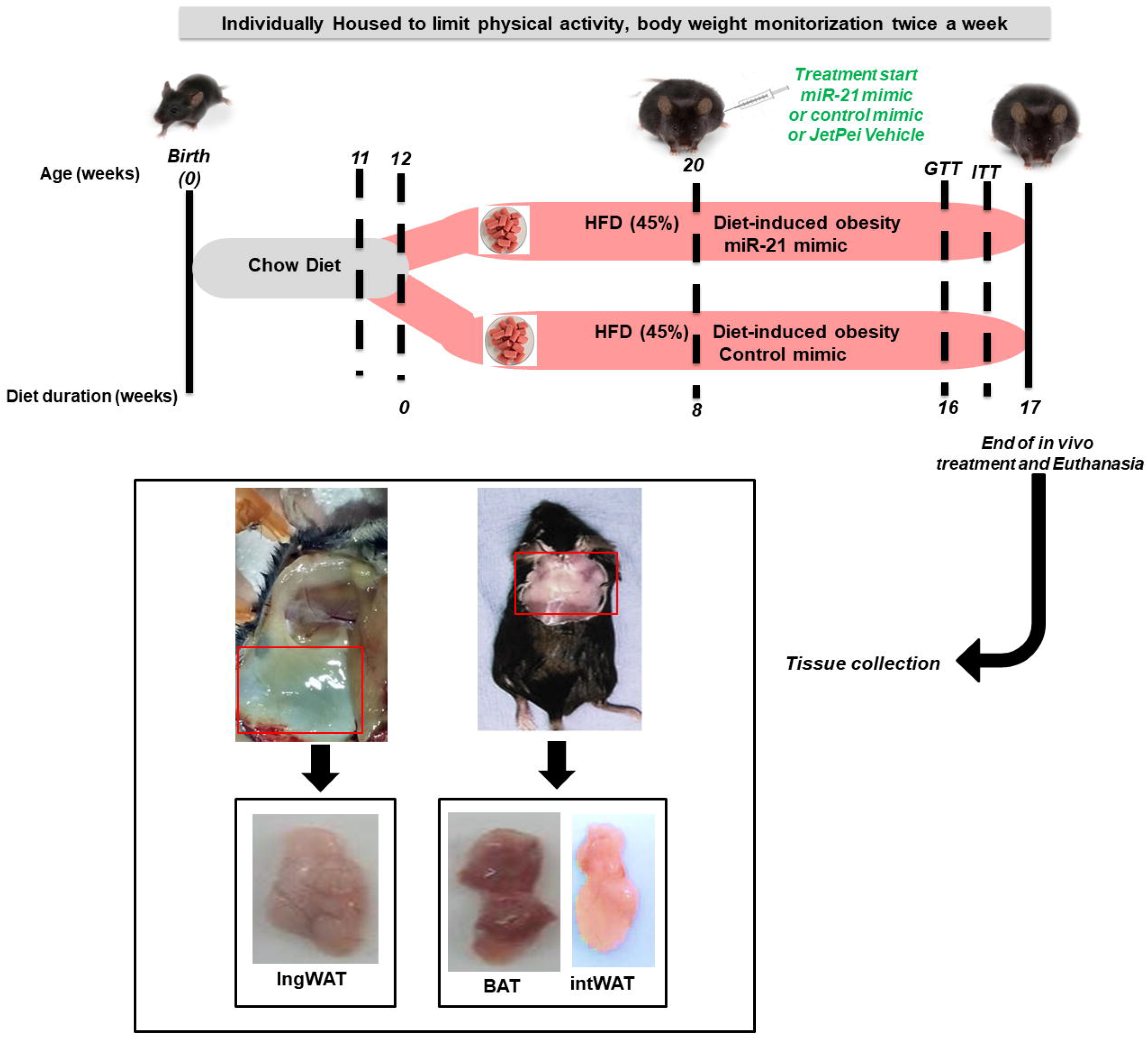
Study design and workflow for *in vivo* miR-21 mimic treatment of HFD-induced obese mice. C57BL/6J mice (11-weeks old at arrival) were fed with chow-diet for one week (quarantine period) and then switched to a high-fat diet (45% kcal from saturated fat) from day 0 (start of the experiment) for 17 weeks. Twice a week, body weight was monitored in awake mice. During these 17 weeks of 45%HFD diet, after the first 8 weeks, mice were injected subcutaneously with miR-21 mimic (0.5 μg, n=8) or control miRNA mimic (0.5 μg, n=9) three times a week for further 8 weeks. Additionally, a subgroup of 45% HFD-mice was injected with jetPEI (0.086 μl, n=4) with the same timing as miR-21 mimic or control mimic (not shown in the graph). At week 16, the glucose tolerance test (GTT) was measured, and two days later, the insulin tolerance test (ITT) was measured. Mice were sacrificed by cervical dislocation at the end of the experiment at week 17. Then, blood samples, interscapular brown (BAT), white interscapular (intWAT) and inguinal (ingWAT) adipose tissues were collected and stored at −80°C until analysis. Mice fed a 45% HFD naive (n=8) and treated with jetPEI (n=4) were also included and weighed (data not shown). The control diet was taken as a reference for body weight gain and glucose tolerance in non-obese mice during the time course of the whole study.

### In vivo sustained treatment with miR-21 mimic decelerated weight gain without affecting glucose and insulin tolerance in HFD obese mice

Given the involvement of miR-21 in the regulation of several genes related to AT function, we explored the effect of miR-21 mimic in obesity. For this purpose, we treated high-fat diet (HFD)-induced obese mice with the miR-21 mimic. To be more precise, the C57Bl/6J strain of mice (susceptible to developing obesity and metabolic disturbances after HFD feeding for several weeks, mimicking human obesity and diabetes (Romero-Zerbo *et al*, 2017)) were used as detailed below. We treated C57BL/6J obese mice (45% HFD) with 0.5 μg of miR-21 mimic or its corresponding miRNA control mimic for an additional period of 8 weeks under HFD. HFD-induced weight and glucose changes previous to miR-21 administration are depicted in **Supplemental Figure S1**. The miR-21 mimic led to a significant deceleration of 45% HFD-induced weight gain compared to the miRNA control mimic (**Fig. 3A**). As illustrated in the AUC weight gain graph, considering the initial weight (prior to miR-21 mimic treatment) as the basal point (0), the mice from each treatment group did not gain weight in the same way: the group treated with miR-21 mimic did not significantly gain weight upon miR-21 administration, while mice treated with the mimic control did. However, no significant amelioration was observed regarding insulin and glucose tolerance with the treatment with the *in vivo* miR-21 mimic (**Fig. 3A**). No changes were observed with vehicle (JetPei) treatment compared to the control (data not shown).

**Figure 3.**
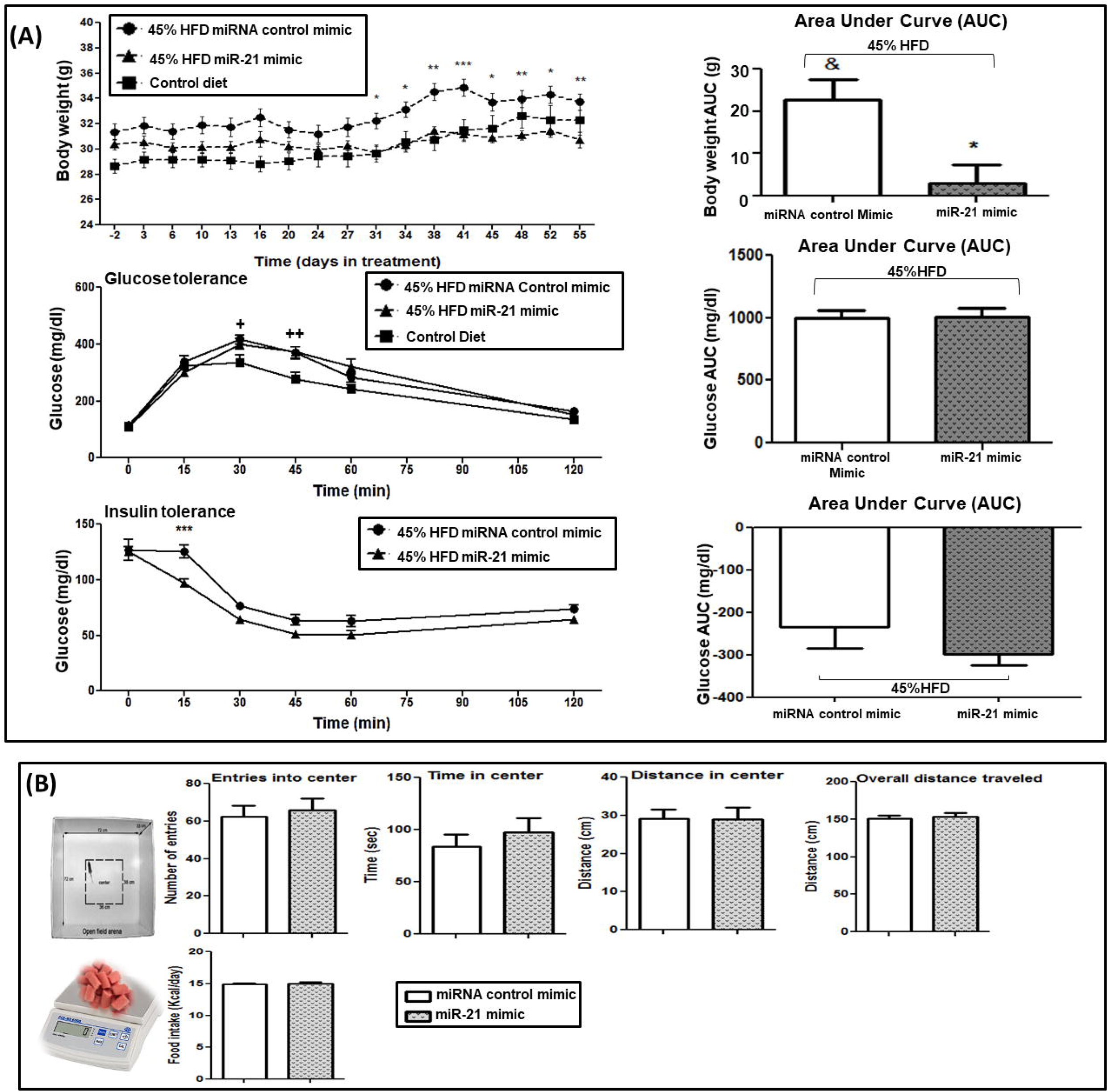
The effects of *in vivo* miR-21 mimic treatment on weight gain, glucose and insulin tolerance tests, food intake and locomotor activity in HFD-induced obese mice. After 8 weeks of 45% HFD, the mice were injected subcutaneously with 0.5 μg of miR-21 mimic or control mimic three times a week for another 8 weeks. **(A)** Body weight was monitored twice a week for the 16 weeks. AUC was calculated using the weight before the start of the treatment or the basal glucose levels before tolerance tests for each mouse as the basal point (0). Glucose (GTT) and insulin (ITT) tolerance tests were carried out during the last week of treatment (week 16) by injecting 2 g/kg of D-glucose or 0.5 U/kg of insulin, respectively, after 10-12 hours of fasting. Blood samples were collected from the tail vein at 0 (basal), 15, 30, 45, 60, and 120 minutes, and glucose measured with a glucometer. **(B) Open field test** (OFT) was performed to measure the locomotor activity of obese mice treated with miR-21 mimic or control mimic. Mice were moved to the experimental room and kept there for 30 minutes before starting the test. Locomotor activity was monitored in the open field arena by video tracking for 10 minutes and the time and distance walked in the center, entries into the center and the overall distance traveled were measured. Five weeks after miR-21 or control mimic treatment, a food intake test was carried out by measuring the mouse body weight and food pellets for 5 days. Daily kcal consumption was calculated based on the high-fat diet. Data are expressed as the mean ± SEM. *p□0.05versus control mimic. ^+^p<0.05 versus control diet. ^++^p<0.01 versus control diet. &p□0.05 versus the basal point (0). Repeated ANOVA measurements and Student’s t-test were used for statistical analysis.

### In vivo effect of sustained treatment with miR-21 mimic on weight loss was independent to locomotor activity or food intake changes in HFD obese mice

To understand whether miR-21-induced weight gain deceleration was due to a change in locomotor activity or food intake, we performed two tests: the open field test and the intake test. The open-field test showed that there were no differences in the number of entrances to the central zone, in the time spent in the central zone, in the distance traveled in the central zone or in the total distance travelled, indicating that no significant differences were recorded in locomotor activity between miR-21 mimic-treated mice and control miRNA mimic-treated mice (**Fig. 3B**). In addition, no significant difference in daily calorie intake was observed between miR-21 mimic- and control mimic-treated mice (**Fig. 3B**).

### miR-21 mimic in vivo treatment-induced thermogenesis and [^18^F]FDG uptake in ingWAT

45% HFD mice both under treatment with miR-21 mimic and control mimic underwent ^18^fluorodeoxyglucose ([^18^F]-FDG) PET-computed tomography (PET-CT). [^18^F]-FDG PET-CT unveiled a significant increase in [^18^F]-FDG uptake in the inguinal WAT (ingWAT) of miR-21 mimic-treated mice compared to control miRNA mimic-treated mice (**Fig. 4A**). As it is well described that subcutaneous WAT adipocytes of rodents are prone to the conversion into thermogenic beige adipocytes and higher expression of *UCP1* and other brown fat cell markers in response to various stimuli, we further examined the expression levels of thermogenic and browning-specific genes in WAT and BAT upon miR-21 treatment. Thermogenic markers (*Pgc-1a, Ucp1, Cidea, Fgf21, Prdm16*) and beige adipocyte-specific markers (*Tmem26* and *Hoxc9*) were analyzed in ingWAT, interscapular WAT (intWAT) and BAT. Consistent with the increased [^18^F]-FDG intake observed in miR-21 mimic-treated mice, *Ucp1, Cidea, Pgc1a, Prdm16*, and *Fgf21* gene expression showed an increase with miR-21 mimic compared to that of the control mimic in both BAT and ingWAT (**Fig. 4B**). Moreover, intWAT displayed specifically increased levels of *Pgc-1a* and *Prdm16* in miR-21 mimic-treated mice compared to levels in control mimic mice. The *Ppar-g* gene expression showed a significant increase in BAT from miR-21 mimic treated mice compared to control mice, while no significant changes were observed in intWAT. Interestingly, the expression of beige adipocyte markers *Hoxc9* and *Tmem26* were significantly higher in ingWAT and intWAT from miR-21 mimic-treated mice compared to control mice.

**Figure 4.**
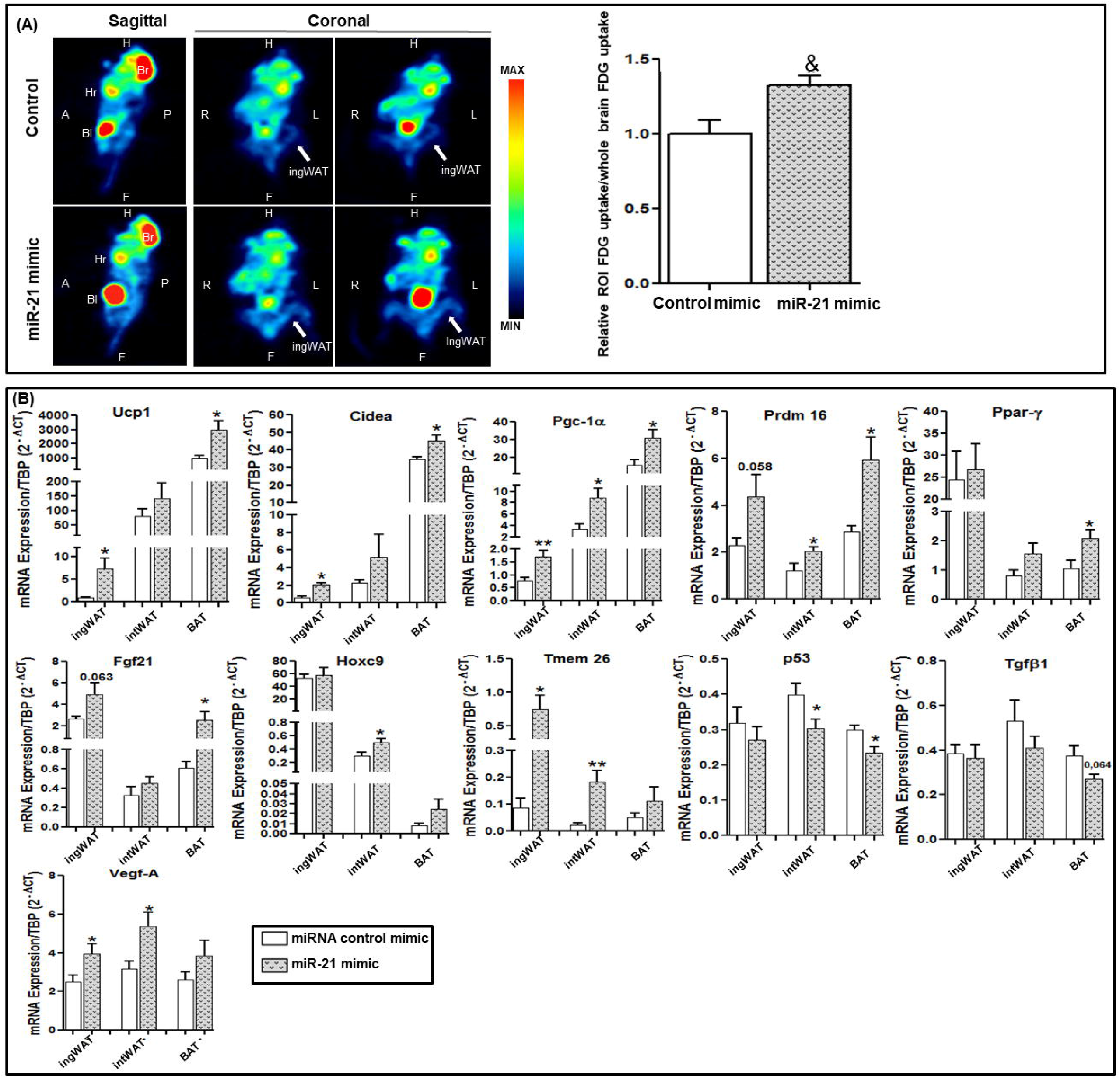
PET analysis of thermogenic activity and browning induction and adipose tissue mRNA expression analysis. **(A)** Quantitative analysis of inguinal white adipose tissue (ingWAT) 18F-Fluorodeoxyglucose ([^18^F]-FDG) uptake imaging from control mimic or miR-21 treated mice was performed by positron emission tomography (PET) (n= 6 per group). [^18^F]-FDG sagittal (left) and coronal (middle and right) PET images of control mimic (top) and miR21 mimic-treated (bottom) mice are shown. The average [^18^F]-FDG uptake was normalized to the whole brain average [^18^F]-FDG uptake. PET data were analyzed using PMOD 3.3 software. Data are expressed as means ± SEM. &p=0.005 according to the paired Student’s t-test). White arrows indicate ingWAT. P: posterior; A: anterior; L: left; R: right; H: head; F: foot; Br: brain; Hr: heart; Bl: bladder. **(B)** mRNA levels of browning, thermoregulatory and angiogenic markers as well as *Tgfβ1* and *p53* from ingWAT, interscapular white adipose tissue (intWAT) and brown adipose tissue (BAT) from mice treated with miR-21 mimic or control mimic (n=7 per group) were measured by real-time qPCR using TATA sequence binding protein (TBP) as a reference gene (2^-Δct^). *p□0.05 versus control mimic. Data are expressed as the mean ± SEM according to the Student’s t-test.

Besides having a relevant role in the modulation of angiogenesis and being a validated gene target of miR-21, VEGF-A is well known to play an important role in the regulation of energetic homeostasis and in obesity and insulin resistance prevention by inducing thermogenesis in BAT and browning activation in WAT (Elias *et al*, 2013; Lu *et al*, 2012) Our data demonstrated that the miR-21 *in vivo* treatment induced an increase in *Vegf-A* mRNA expression levels in both ingWAT and intWAT tissues compared to that in the control treatment (**Fig. 4B**).

On the other hand, the gene expression of *Tgfβ1* and *p53* (reported to inhibit the differentiation and thermogenesis in BAT, to regulate the formation of beige cells in WAT, and to be associated with obesogenic pathways, T2D and IR (Hallenborg *et al*, 2016; Yadav *et al*, 2011)) was significantly downregulated with miR-21 *in vivo* treatment compared to that in control mimic mice (**Fig. 4B**).

In order to ascertain whether *in vivo* miR-21 mimic treatment increased miR-21 expression levels in the tissues of interest, the expression levels of miR-21 were analyzed in VAT, ingWAT, BAT and plasma from miR-21 mimic and control groups. **Figure S2** shows an increase in miR-21 levels with *in vivo* miR-21 mimic treatment, significantly so in VAT, ingWAT and BAT, compared to controls, confirming that miR-21 treatment led to an increase in the levels of this miRNA in the analyzed tissues.

### miR-21 mimic in vivo treatment induced the appearance of brown-like adipocytes with numerous small lipid droplets and mitochondria in WAT

Hematoxylin-eosin staining revealed that the intWAT and ingWAT of miR-21 mimic *in vivo-*treated mice accumulated a large number of multilocular adipocytes, while the intWAT and ingWAT from control miRNA mimic-treated mice had mostly large adipocytes with unilocular lipid inclusions (**Fig. 5A and 5B**). Moreover, immunofluorescence staining showed that intWAT and ingWAT from miR-21 mimic *in vivo*-treated mice displayed a much higher UCP1 and TMEM26 protein expression compared to control mimic. The merged image demonstrated the co-expression of both TMEM26 and UCP1 in both ingWAT and intWAT from miR-21 mimic-treated mice and control mice. In BAT from miR-21 mimic*-*treated mice, multilocular adipocytes were present. In addition, immunostaining showed that UCP1 displayed a clear expression signal in both groups of mice, while no signal corresponding to TMEM26 was detected (**Fig. 5C**). Moreover, transmission electron microscopy (TEM) was used to compare the ultrastructure of the multilocular adipocytes and their mitochondria from miR-21 mimic ingWAT with that from control miRNA mimic ingWAT. **Fig. 5D** shows the presence of numerous mitochondria that are larger in ingWAT from miR-21 mimic-treated mice compared to control mice.

**Figure 5.**
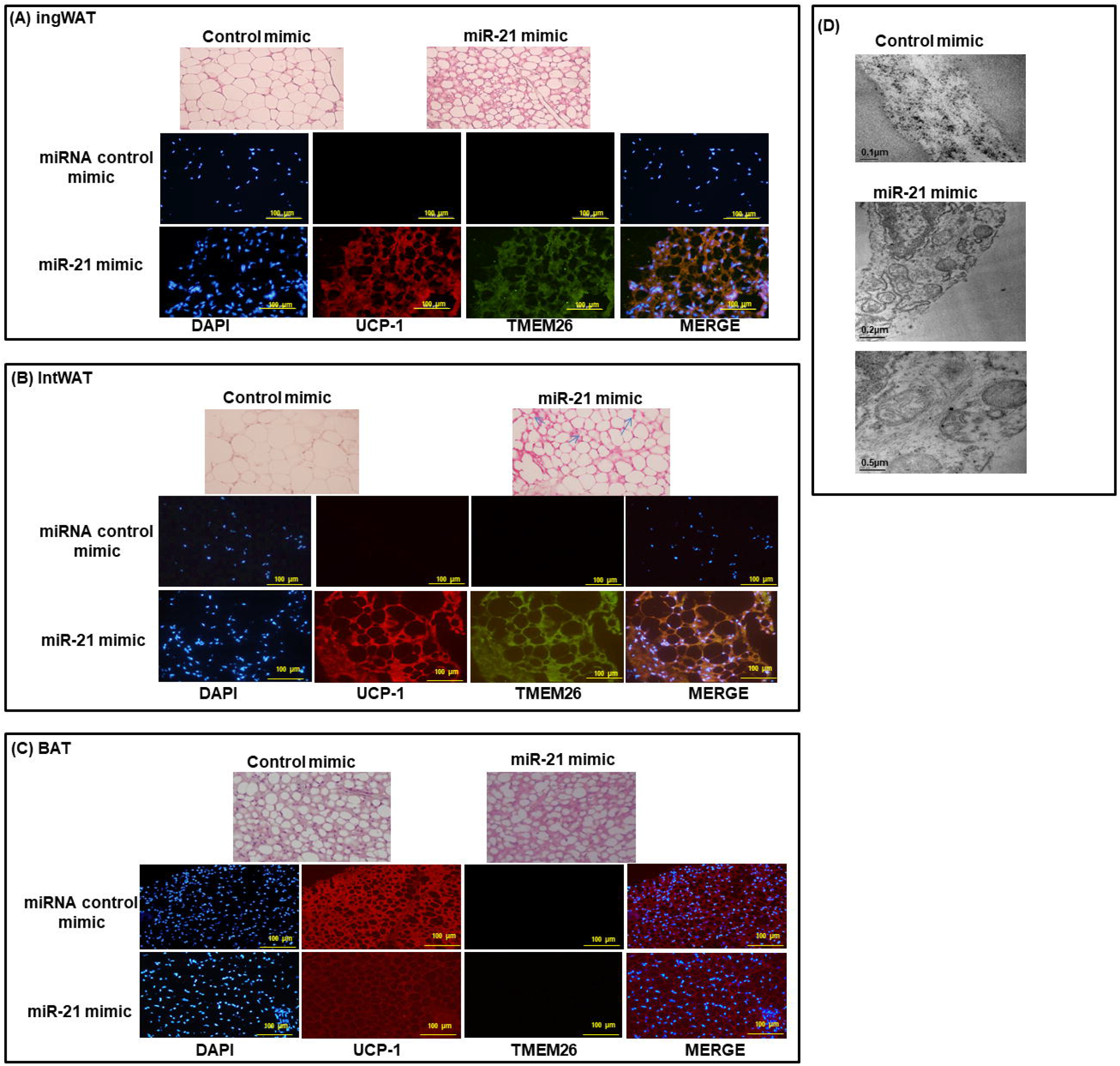
Histological analysis of WAT and BAT, confocal imaging of browning and thermogenic proteins and transmission electron microscopy (TEM) of white adipocytes. After *in vivo* treatment with miR-21 mimic or control mimic, inguinal white adipose tissue (ingWAT) **(A)**, interscapular white adipose tissue (intWAT) **(B)** and brown adipose tissue (BAT) **(C)** samples were stained by hematoxylin and eosin (X20). IngWAT **(A)**, intWAT **(B)** and BAT **(C)** were immunostained with goat anti-UCP1 (red), rabbit anti-TMEM26 (green) and nuclei stained with DAPI (blue). Images were visualized by confocal microscopy (X20). **(D)** Transmission electron microscopy (TEM) images of cell organelles from ingWAT of mice treated with miR-21 mimic or control mimic. The micrographs are presented with scale bars of 0.5 μm and 0.2 μm.

### miR-21 mimic in vivo treatment increased mtDNA copy number and Sirtuin 1 mRNA expression levels in inguinal WAT

We further confirmed the increase in the number of mitochondria in ingWAT from miR-21 mimic treated mice compared to controls by quantifying the copy number of the mitochondrial DNA (mtDNA), which reflects the abundance of mitochondria within a cell (**Fig 6A**).

**Figure 6.**
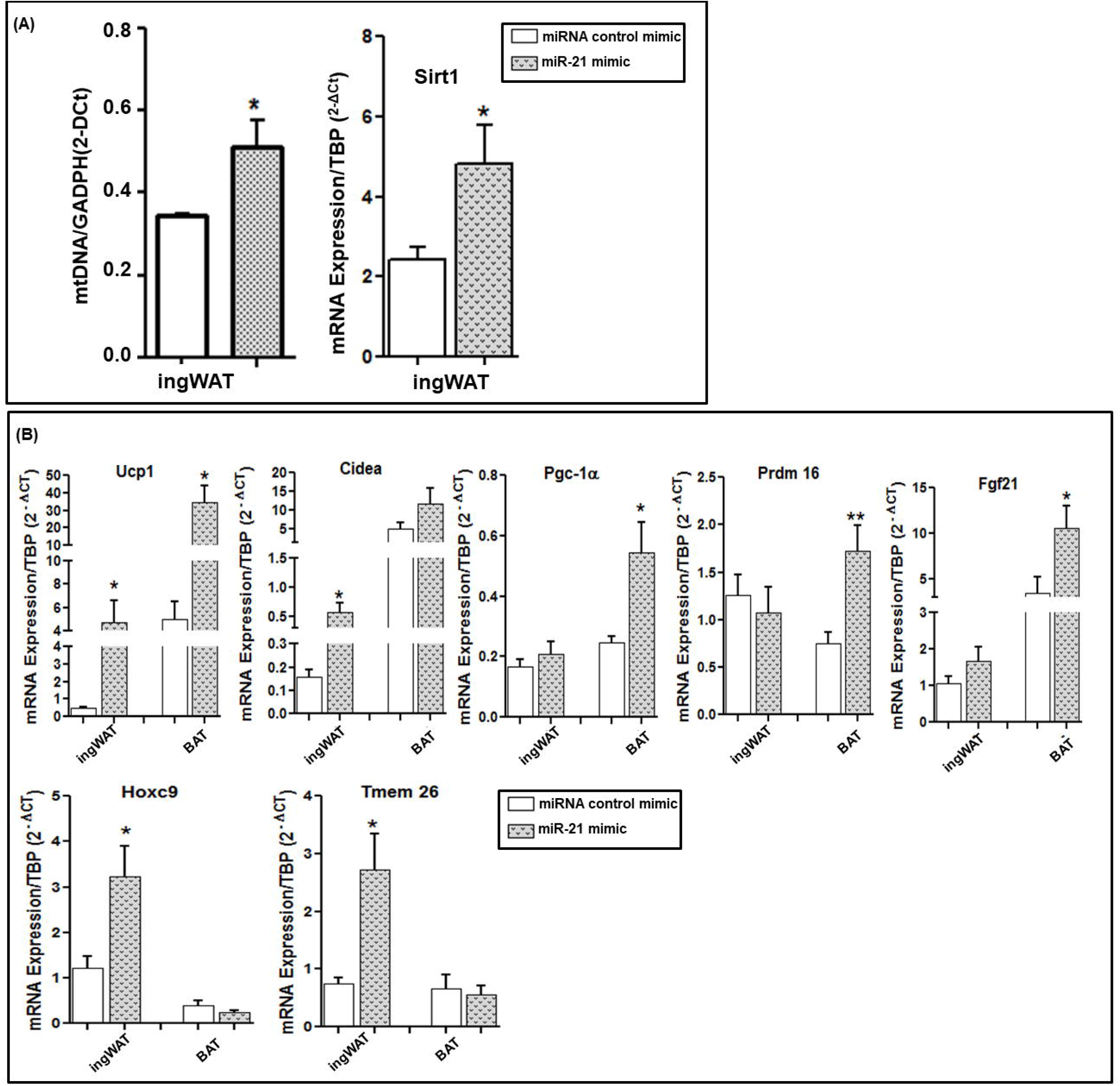
*In vivo* effect of miR-21 mimic on mtDNA and *Sirtuin 1* expression levels and *in vitro* effect of miR-21 mimic on the mRNA expression of browning and thermogenesis markers of adipose tissue explants. **(A)** After *in vivo* treatment with miR-21 mimic or control mimic, COX1 as mitochondrial gen and GAPDH were measured by real-time qPCR and the relative mtDNA content was calculated the formula: mtDNA= 2^-Δct^, where ΔCt= Ct COX1-Ct GADPH. *Sirt1* mRNA expression was measured from the inguinal white adipose tissue (ingWAT) of mice treated with miR-21 mimic or control mimic by real-time qPCR using TATA sequence binding protein (TBP) as a reference gene (2^-Δct^). Data are expressed as the mean ± SEM. *p□0.05 versus control mimic a control Student’s t-test was used for statistical analysis. (**B**) Fresh BAT and ingWAT explants from mice fed a control diet (10% kcal fat) were incubated for 48 hours at 37°C with 5 nM miR-21 mimic or control mimic (n=7). The mRNA expression of browning and thermoregulatory markers was measured by real-time qPCR using TATA sequence binding protein (TBP) as a reference gene (2^-Δct^). Data are expressed as the mean ± SEM. *p□0.05 and **p□0.01 versus control mimic according to the Student’s t-test.

We also analyzed *Sirt1* mRNA expression as it is well known that Sirt1 is a protein that plays a pivotal role in promoting mitochondrial biogenesis and metabolic control by means PGC-1*α* deacetylation (Tang, 2016). In agreement with our mtDNA copy number and microscopy results, *in vivo* miR-21 mimic treatment induced a significant increase in *Sirt1* expression levels in ingWAT compared to control mimic (**Fig. 6A**).

### In vitro treatment with miR-21 mimic induced thermogenic and browning gene expression in ingWAT and intBAT mice explants

The effects of miR-21 mimic were analyzed *in vitro* in AT explants from mice fed with control diet (**Fig. 6B**). IngWAT explants *in vitro* treated with miR-21 mimic displayed a significant increase in *Ucp1, Cidea, Hoxc9* and *Tmem26* gene expression compared to explants treated with control mimic. A significant increase in *Ucp1* and *Prdm16* gene expression in BAT explants upon miR-21 mimic treatment compared to control mimic was also detected, confirming thermogenesis activation and browning induction seen with the *in vivo* miR-21 treatment in mice.

### The effect of miR-21 on browning in WAT could be through the signaling pathways mediated by Vegf-A, Pgc1a, Prdm16, Pparγ, Fgf21, Tgfβ1, and p53 genes, which lead to Ucp1, Cidea, and Tmem26 gene transcription induction

As described above, we have experimentally shown that *Pgc-1α, Vegf-A, Tgf-β1* and *p53* are potential target genes for miR-21 (**Fig. 7A**). Up to date, *Pparγ, Prdm16* and *Tmem26* genes were predicted, but not validated, miR-21 target genes according to our *in-silico* analyses (**Supplemental Tables 1 and 2)**. PANTHER and GeneCodis3 showed that these target genes are mostly involved in thermogenesis and browning processes (**Fig. 7A**). This work demonstrated that *Ucp1, Hoxc9, Cidea*, and *Fgf21* genes could be potential key regulators for thermogenesis and browning (**Fig. 7A**). In addition, the inhibition of *Tgfβ1* and *p53* could also be another miR-21 regulated pathway by which this miRNA regulates WAT function (**Fig. 7B**).

**Figure 7.**
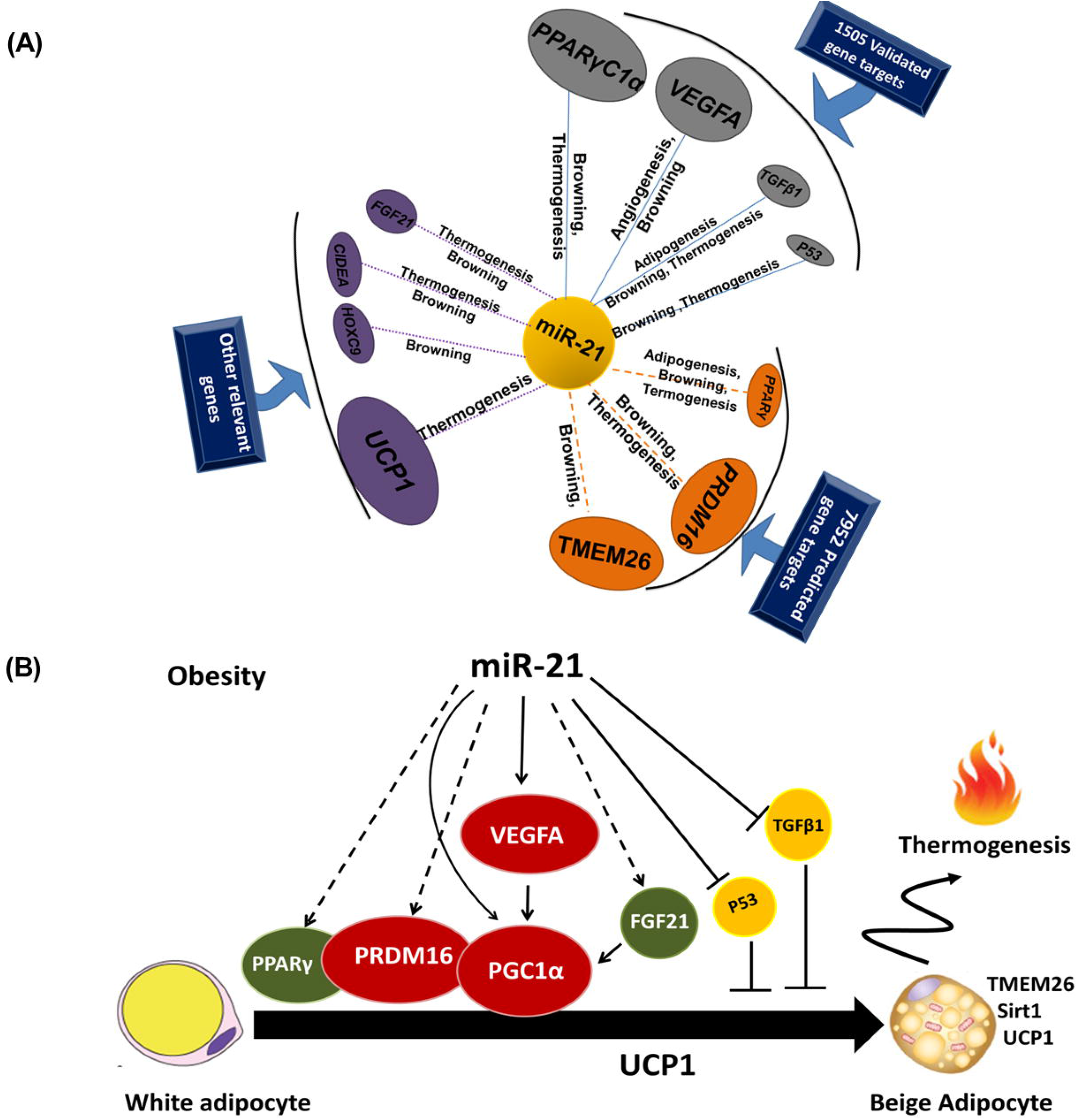
Schematic potential target genes and model pathways that might be involved in the miR-21 effects in AT. **(A)** Schema summarizing the interactions among miR-21, biologic processes, and the target genes. The validated and predicted target genes of miR-21 were obtained using miRTarBase 4.0, Tarbase v8 and miRWalk 2.0. The PANTHER Classification System and GeneCodis3 (Gene annotations co-occurrence discovery) were applied to annotate the biological processes of the predicted targets. The interactions among miR-21, biologic processes, and the target genes were visualized with Cytoscape version 3.2.1 software. **(B)** Scheme showing the possible signaling pathways by which miR-21 could act in the activation of thermogenesis and browning of WAT of mice.

TargetScan tool analysis, which allows us to predict the binding sites of miRNA within the objective (Shao, 2017), showed that miR-21 has effective binding sites only within *Ucp1* (context ++ score percentile: 97%), with a P_CT_ value that indicates the probability of segmentation being conserved for a single target site, showing values of <0.1, indicating an elevated evolutionary conservation (**Table S3**).

### Visceral and subcutaneous WAT miR-21 expression is enhanced in obese patients and mice without T2D or insulin resistance

In order to analyze whether miR-21 overexpression could be related to the maintenance of metabolic health despite the excess weight, we aimed at analyzing WAT miR-21 levels regarding this phenotype. VAT and SAT miR-21 expression levels were significantly higher in obese subjects with a low degree of insulin resistance (LIR-MO) compared to healthy NW individuals. Although a moderate downward trend was seen when compared with LIR-MO, no significant differences were detected in VAT or SAT from obese subjects with a high degree of insulin resistance (HIR-MO) in comparison with healthy NW or LIR-MO (**Fig. 8A**).

**Figure 8.**
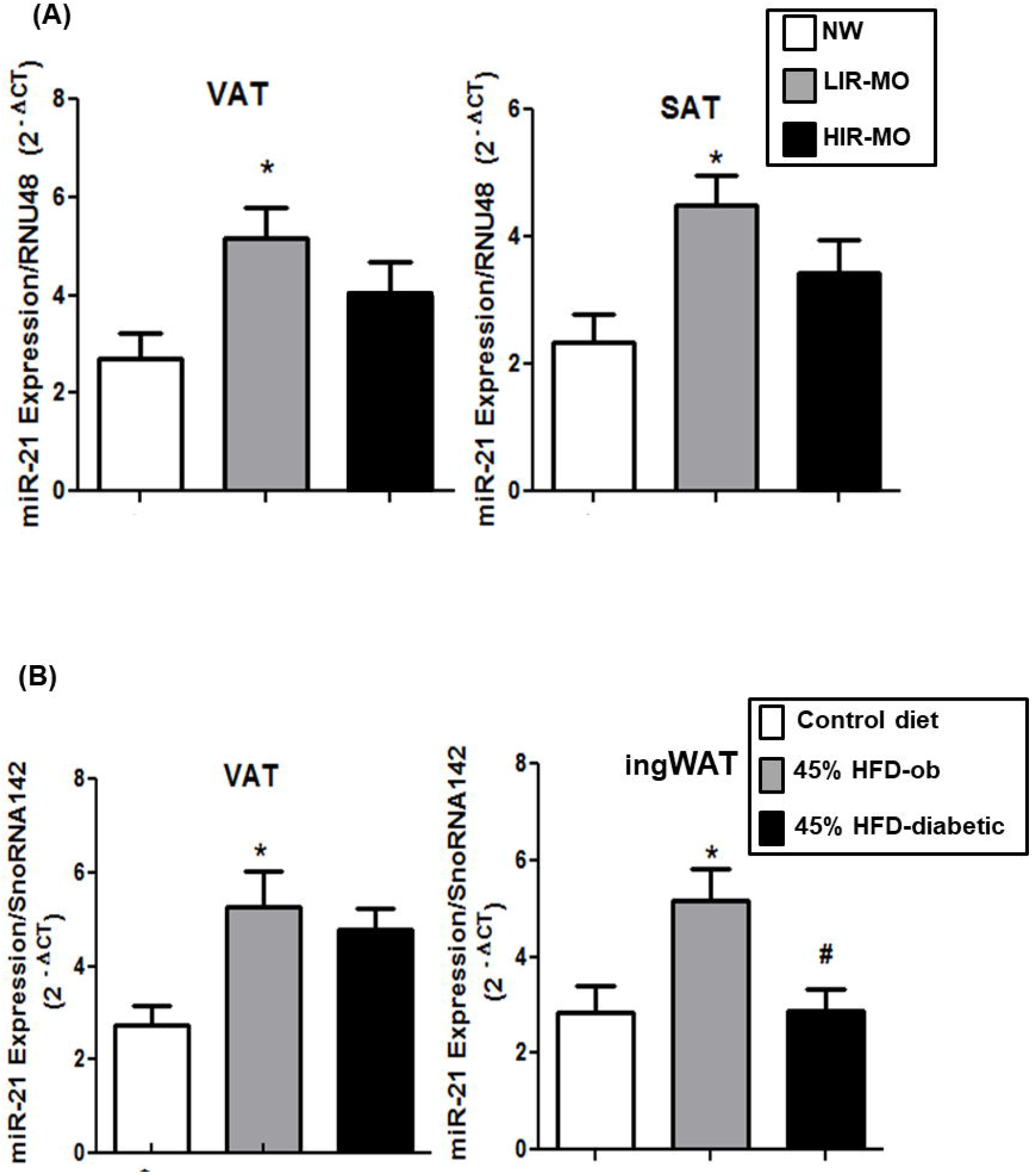
miR-21 expression levels in VAT and SAT from humans and mice. miR-21 expression was measured in **(A)** VAT and SAT from non-diabetic normoweight subjects (NW, n=7), low insulin-resistant morbidly obese subjects (LIR-MO, n=9), high insulin-resistant morbidly obese subjects (HIR-MO, n=9), and **(B)** in ingWAT and VAT from C57BL/6J mice fed a low-fat diet (10% kcal fat; control diet; n=9), non-diabetic obese mice fed a high-fat diet (45% kcal fat) during 8 weeks (45% HFD-ob; n=8) and diabetic obese fed a high-fat diet (45% kcal fat) during 14 weeks (45% HFD-diabetic; n=8), by real-time qPCR using RNU48 (human) and SnoRNA142 (mice) as reference genes to calculate 2^-Δct^ according to the manufacturer indications. Data are expressed as the mean ± SEM. *p□0.05 versus NW or control; #p□0.05 versus 45% HFD-ob according to One-way ANOVA with Bonferroni’s correction.

The C57Bl/6J strain of mice (susceptible to developing obesity and metabolic disturbances after HFD feeding for several weeks) was used to mimic human obesity and diabetes (Romero-Zerbo *et al*, 2017). The study was carried out in three groups of mice: mice fed with control diet (control), non-diabetic obese mice (45% HFD for 8 weeks; 45% HFD-ob) and obese diabetic mice (45% HFD for 14 weeks; 45% HFD-diabetic) (**Fig. S3A**). Between 34-51 days, the body weight gain of both HFD-ob and HFD-diabetic groups began to be significantly greater than that of control mice (**Fig. S3B**). At the end of the study, the HFD-diabetic group showed significant glucose and insulin intolerance compared to control group (**Fig. S3C**). Similar to human results, miR-21 expression levels in VAT and ingWAT from 45% HFD-ob mice were significantly higher compared to control mice. Notably, miR-21 levels in ingWAT from 45% HFD-ob were significantly higher compared to those in 45% HFD-diabetic mice (**Fig. 8B**).

## Discussion

We here describe evidence showing that miR-21 is involved in the regulation of AT functionality, by controlling a number of genes and biological processes including thermogenesis and browning, angiogenesis, VEGF signaling, apoptosis and adipogenesis, suggesting the pivotal role that miR-21 could be playing in obesity and its related metabolic alterations such as T2D and IR, by modulating AT physiology.

While investigating the mechanisms underlying the effects of the previously reported increase in WAT miR-21 levels in obesity and T2D(Guglielmi *et al*, 2017; Keller *et al*, 2011), we unexpectedly realized that the mimetization of miR-21 may be a potential effective therapy for obesity. Specifically, the administration of miR-21 mimic in obese mice stopped the HFD-induced weight gain independently of food intake or physical activity. Interestingly, these were accompanied by the activation of browning and the thermogenic programming in WAT corroborated by changes in metabolic activity, protein expression, and cellular ultrastructure. This was further supported by the upregulation of browning and thermogenesis markers in WAT and BAT. Concordant with the apparent protective role against obesity, miR-21 high expression levels were associated with metabolically healthy obesity in humans and mice. These data establish miR-21 mimic as a potential therapy for obesity by inhibiting weight gain in a healthy manner.

It is well described that deregulation of WAT apoptosis, adipogenesis and angiogenesis are described to underlie AT dysfunction in obesity, which in turn increases the risk of T2D and IR(Blüher, 2009). Here, miR-21 mimic treatment seems to regulate WAT angiogenesis through increasing the expression levels of the pro-angiogenic factors *Vegf-A* and *Vegf-B. Vegf-A* is a key regulator of angiogenesis, a validated target gene of miR-21 and its overexpression in AT has been reported to protect against diet-induced obesity and IR (Elias *et al*, 2012). *Vegf-B* has also been associated with an enhanced insulin delivery and function in AT, resulting in an improvement of metabolic health in obesity (Karaman *et al*, 2016). In agreement with the pro-angiogenic role of miR-21, it has been previously reported that miR-21 triggers angiogenic mediators, such as *Timp3, Ang2* and *Ang4*, in several cell types. These factors have been involved in regulating angiogenesis in WAT and are associated with positive metabolic features (Hu *et al*, 2016; An *et al*, 2017).

We also saw that miR-21 mimic upregulates *Bcl2, Bid* and *Casp3* genes in adipocytes, which have been well described to play a pivotal role in controlling the impairment of insulin signaling in human AT through the regulation of the inflammatory processes and the development of the obesity-associated insulin resistance (Alkhouri *et al*, 2010; Tinahones *et al*, 2013). Surprisingly, in adipocytes, miR-21 mimic appears to significantly reduce the expression levels of both *Cebp-α* and *Ppar-γ*, which are the master transcriptional regulators of adipogenesis (White & Stephens, 2010), while increasing the expression of *Pgc-1α* and *Fgf21*, genes well known to play a relevant role in BAT thermogenesis and WAT browning (Coskun *et al*, 2008; Fisher *et al*, 2012). Altogether, these results suggest that miR-21 mimic treatment, as well as regulating the key mediators of WAT functionality and expansion, also seems to promote the switch to the thermogenic adipocyte cell lineage while reducing the process related to white adipocytes cell lineage.

Interestingly, sustained exogenous administration of miR-21 mimic in obese mice abolished HFD-induced weight gain. It is well known that obesity and body weight are associated with the energy balance between food intake and energy expenditure (Slawik & Vidal-Puig, 2007). Within this context, the fact that the *in vivo* miR-21 mimic treatment did not affect locomotor activity or calorie intake compared to controls, suggests a direct involvement of miR-21 in mechanisms regulating fat accumulation related to the activation of thermogenesis and browning, two processes that have arisen as potential targets for the management of obesity and its related diseases (Peirce *et al*, 2014). Several studies have shown that the activity of brown and beige adipocytes is associated with resistance to obesity in several mouse models (Cederberg *et al*, 2001; Kopecky *et al*, 1995; Seale *et al*, 2008). In addition, it is well recognized that increased activity of brown and beige fat not only affects the weight gain of mice but also improves systemic metabolism, including the improvement in glucose tolerance and insulin sensitivity (Cederberg *et al*, 2001; Seale *et al*, 2008; Bordicchia *et al*, 2012; Boström *et al*, 2012). Many miRNAs have been reported to be related to the regulation of genes mediators of beige and brown adipocytes differentiation(Mori *et al*, 2012; Sun *et al*, 2011; Trajkovski *et al*, 2012). However, the involvement of miR-21 in the regulation of thermogenesis and browning gene expression has not yet been previously described. Here, we show that miR-21 mimic could play a pivotal role in obesity and weight gain control through browning and thermogenesis activation involving the induction of *Tmem26, Pgc-1α, Ucp1, Prdm 16, Fgf21, Cidea, Pparγ and Vegf-A* gene overexpression and mitochondrial biogenesis process.

Our *in-silico* prediction study identified validated miR-21 target genes, including *Vegf-A, Ppargc-1a, Tgfβ1* and *p53*. These genes are primarily involved in the regulation of browning thermogenesis, adipogenesis, and angiogenesis (Elias *et al*, 2013; Hallenborg *et al*, 2016; Yadav *et al*, 2011), as well as predicted (non-validated) miR-21 target genes, including *Ppar-γ, Prdm16* and *Tmem26*, which are primarily known to be key regulators of browning and thermogenesis in AT (Seale *et al*, 2011, 2008; Petrovic *et al*, 2010) (**Fig. 7A**). We also provided experimental evidence through our *in vivo, ex vivo* and *in vitro* studies of novel gene targets of miR-21 in WAT, namely, *Ucp1, Hoxc9, Cidea* and *Fgf21*. Interestingly, the TargetScan tool identified miR-21 binding sites within *Ucp1* mRNA. Further experiments should be carried out to ascertain whether *Ucp1, Hoxc9, Cidea* and *Fgf21* are direct target genes of miR-21.

Our comprehensive study allowed us to build a potential regulatory model that proposed that miR-21 regulates browning and thermogenesis through several ways: 1) VEGF-A signaling pathway, 2) p53 signaling pathway and 3) TGFβ1 pathway (**Fig. 7B**). In the line with our suggestions, *Vegf-A* has been described to be a miR-21 target gene (Lei *et al*, 2009; Liu *et al*, 2011) and has also been described to induce the expression of *Pgc-1α* and *Ucp1* in both mouse BAT and WAT (Elias *et al*, 2013) and then, this factor regulates energy homeostasis and has a protective effect against obesity and IR, not only by angiogenesis regulation but also through the induction of BAT thermogenesis and WAT browning (Elias *et al*, 2013; Lu *et al*, 2012). As illustrated in **Figure 7B**, the activation of *Ucp1* by miR-21 could also occur through pathways involving *Fgf21, Prdm16* and *Ppar-γ*. In fact, *Fgf21* has been well described to induce thermogenesis in BAT and browning in WAT by activating *Pgc-1α* (Coskun *et al*, 2008; Fisher *et al*, 2012). *Pgc-1α* and *Prdm16* have also been shown to induce the expression of *Ucp1* and other thermogenic components and to regulate energy homeostasis and obesity (Puigserver *et al*, 1998; Tiraby *et al*, 2003; Villanueva *et al*, 2013). Moreover, miR-21 could act on browning and thermogenic processes through the inhibition of *p53* and *Tgfβ1*, two validated miR-21target genes that were downregulated in both the *in vivo* and *in vitro* miR-21 mimic treatment. These two target genes have been well described to inhibit differentiation and thermogenesis in BAT, to regulate the formation of beige cells in WAT and to be related to obesity, diabetes and insulin resistance (Yadav *et al*, 2011; Hallenborg *et al*, 2016).

Altogether, our data suggest the benefits of increasing miR-21 levels in AT as a potential therapy against weight gain which likely prevents the switch to an unhealthy metabolic phenotype. In this regard, we found that high levels of miR-21 in both VAT and SAT were associated with metabolically healthy obesity defined by a low degree of insulin resistance in humans and the absence of T2D in mice. Consistent with our findings, previous studies reported that miR-21 is highly expressed in human SAT, and correlated with BMI (Keller *et al*, 2011). In addition, an increase in miR-21 has been observed in the VAT of HFD-fed C57BL/6J mice compared to that of low-fat diet-fed mice (Chartoumpekis *et al*, 2012). Overall, our human and mice transversal studies support the potential role of miR-21 exogenous increase for the regulation of WAT functionality and expansion and suggest that high levels of miR-21 in AT could be a protective mechanism that allows obese subjects to remain metabolically healthy. This hypothesis is in agreement with our previous results which pointed out the upregulation of VEGF-A expression as a defense mechanism against obesity-associated metabolic alterations such as IR and T2D (Tinahones *et al*, 2012). Here we can add that this VEGF-A increase could be triggered by miR-21, as both *in vitro* and *in vivo* miR-21 mimic administration led to an increase in *Vegf-A* gene expression. Future analyses should be carried out to address these hypotheses.

In conclusion, to the best of our knowledge, this study adds novel findings that highlight the role of miR-21 as a promising potential therapeutic strategy for limiting weight gain and maintaining metabolic health.

## Materials and Methods

### Effect of miR-21 on differentiated 3T3-L1 adipocytes

3T3-L1 cells (ATCC-CL-173) were cultured in DMEM/F12/10% FCS at 37°C, 95% humidity and 5% CO_2_ (n=6). Differentiation of 3T3-L1 was induced by incubation in DMEM/F12/10% FCS supplemented with 0.5 mM isobutylmethylxanthine, 1 μM dexamethasone and 1.67μM insulin for 48 hours and then in DMEM/F12/10% FCS supplemented with 1.67μM insulin for 5 days. Seven days after adipogenic induction, the adipocytes were incubated with 5nM miR-21 mimic or control mimic for 48 hours. Dharma FECT Transfection Reagent was used to transfect cells.

### Study design for *In vivo* treatment of 45% HFD fed mice with miR-21 mimic

To obtain obese mice, a group of mice were fed a HFD containing 45% of kcal from saturated fat for 8 weeks (45% HFD). Body weight was monitored twice a week. After 8 weeks of feeding, glucose tolerance was assessed by intraperitoneal glucose tolerance test (GTT) (Fig. S1). Body weight and glucose areas under the curves (AUCs) were calculated using the initial weight before starting DIO and basal glucose after 10-12 hours of fasting as the basal point (0) for each mouse (Fig. S1). In this step of the *in vivo* study, the 45% HFD mice group was compared in parallel with a control group of age-matched mice which were fed a control diet containing 10% of kcal from fat (LFD) in order to ensure that the 45% HFD mice group had significant differences in weight and glucose tolerance compared to 10% LFD group (control diet) after 8 weeks of diet, which indicates that miR-21 *in vivo* treatment can be started. The control diet group was then discarded, as the study was focused mainly on obese mice. The obese mice were separated into three subgroups: 1) a group treated three times a week for 8 weeks with subcutaneous injection of 0.5μg miR-21 mimic (n=8) (Riboxx, M-00303-0100) dissolved in jetPEI vehicle (0.08μl) (polyplus transfection, 201-50G); 2) a group treated three times a week for 8 weeks with mimic control (n=9) (Qiagen, AllStars Neg. Control siRNA) dissolved in jetPEI vehicle; and 3) a group treated three times a week for 8 weeks with jetPEI vehicle (n=4). All animals were sacrificed by cervical dislocation. Blood and AT biopsies, BAT, interscapular white AT (intWAT), inguinal AT (ingWAT) and visceral AT (VAT), were obtained during dissection and stored at −80°C. The number of mice in each experiment is stated in the figure legends (Fig. 2).

### Participants for the human study on miR-21 expression

Obese subjects were included in this study based on the inclusion criterion of morbid obesity (BMI[40) within two groups. The first group contains subjects who are morbidly obese with low insulin resistance (homeostasis model assessment of insulin resistance index (HOMA-IR) <3.55) (LIR-MO, n=9). The second group contains subjects who are morbidly obese with high insulin resistance (HOMA-IR>6.73) (HIR-MO, n=9). The morbidly obese subjects included in this study (n=18) received bariatric surgery at the Hospital Clínico Virgen de la Victoria. Control subjects were non-diabetic normoweight individuals with HOMA-IR< 3.55 (NW) (BMI=18.5-24 kg/m^2^, n=7) who were operated for hiatal hernia or cholelithiasis, age-matched to the two morbidly obese groups, and with the same inclusion criteria. The exclusion criteria were (1) T2D with medical treatment, (2) major cardiovascular disease in the 6 months prior to inclusion in the study, (3) evidence of acute or chronic inflammatory disease, (4) infectious diseases, and (5) refusal of the patient to participate in the study. The experimental protocol was approved by the Ethics and Research Committee of the Vírgen de la Victoria Clinical University Hospital (Málaga, Spain). The biochemical and anthropometric characteristics of these subjects are shown in Table S4. The anthropometric measurements of the morbidly obese subjects (LIR-MO and HIR-MO) demonstrated a significantly higher BMI and waist circumference compared to those of the NW subjects (Table S4). The control normoweight subjects had no alterations in their lipid or glucose metabolism, were of a similar age as the morbidly obese group and reported that their body weight had been stable for at least 3 months prior to the study. The biochemical abnormalities associated with obesity and IR were reflected by low levels of HDL cholesterol in both LIR-MO and HIR-MO subjects compared to that of NW subjects. No differences were observed in the HOMA-IR between LIR-MO subjects and NW, while a significant difference in this index was observed between LIR-MO and HIR-MO. AT biopsies were obtained from subcutaneous and visceral (omental) areas during surgery and were frozen and stored at −80°C for posterior miRNA extraction.

### Generation of diet-induced obese and diabetic mice and study design for miR-21 expression studies

The C57Bl/6J strain of mice used in the present study is susceptible to developing obesity and metabolic disturbances after being fed a high-fat diet (HFD) for several weeks (Romero-Zerbo *et al*, 2017). As we previously described, this mouse strain on a HFD for 10 weeks led to the development of a phenotype mimicking that of human obesity and diabetes (Romero-Zerbo *et al*, 2017). Briefly, C57BL/6J mice (11 weeks old at arrival) were purchased from Charles River, France, and were allowed to acclimatize in the animal facility for one week prior to the experiments. Mice were singly housed under a 12-h light/dark cycle (8:00□pm lights off) in a room with controlled temperature (21□±□2°C) and humidity (50□±□10%) and free access to pelleted chow (Standard Rodent Diet A04, SAFE, Panlab, Barcelona, Spain) and water. The study was carried out in three groups of mice: mice fed with control diet (control), non-diabetic obese mice (45% HFD for 8 weeks; 45% HFD-ob) and obese diabetic mice (45% HFD for 14 weeks; 45% HFD-diabetic) (**Fig. S1A**). Specifically, to obtain obese non-diabetic mice (45% HFD-ob), a group of eight mice were fed a control diet containing 10% of kcal (control diet) (D12450 Research Diets Inc) for 6 weeks and then were fed, for further 8 weeks, a HFD (D12451 Research Diets Inc, New Brunswick, NJ, USA) containing 45% of Kcal from saturated fat (HFD (45% HFD). To obtain diabetic obese mice a group of eight mice were fed a HFD containing 45% of kcal from saturated fat for 14 weeks (45% HFD-diabetic). A control group of mice were fed a control diet containing 10% of Kcal from fat (control) for the 14 weeks (**Fig. S3A**). Twice a week, body weight was monitored. As previously described (Romero-Zerbo *et al*, 2017), at week 13, the 45% HFD mice group displayed a significant difference in body weight gain compared to control diet group (**Fig. S3B**). After 14 weeks of feeding, glucose and insulin tolerance were assessed by intraperitoneal glucose tolerance test (GTT and ITT) (**Fig. S3C**). Then, mice were euthanized by cervical dislocation. Moreover, the group of diabetic obese mice showed significant alterations in GTT and ITT compared to the control diet group (**Fig. S3C**). Tissues were immediately collected for further histological and biochemical analyses. The European Union recommendations (2010/63/EU) on animal experimentation were followed. All procedures were approved by the ethics committee of the University of Malaga (authorization no. 2012–0061A).

### In vitro treatment of explants with miR-21 mimic

For the *ex vivo* study, fresh inguinal subcutaneous and brown AT explants (50 mg, n=7) from mice fed a control diet (Low-fat diet, 10% kcal from fat) were dissected and cut into small pieces (5mg-10mg). All explants were preincubated with PBS supplemented with 5% BSA for 30 minutes and then in 199 medium supplemented with 10% FBS, 100 units/mL penicillin and 100 μg/mL streptomycin for 1 hour at 37°C. Then, 5 nM miR-21 mimic or control mimic was added, and explants were incubated at 37°C for 48 hours. Dharma FECT Transfection Reagent was used to transfect AT explants.

### Glucose and insulin tolerance tests

Both the glucose tolerance test (GTT) and insulin tolerance test (ITT) were assessed after eight weeks and 17 weeks of diet (control diet and diet-induced obesity) in the generation of diet-induced obese mice (Fig. S3). In the *in vivo* study, GTT was assessed in the mice groups treatment at week 8 (before the start of treatment) and at the end of treatment study (Fig 3A and S1).

Mice were injected intraperitoneally with 2 g/kg of D-glucose (Sigma-Aldrich, St. Louis, MO) after 10-12 hours of fasting before starting the treatment with miR-21 mimic and at the end of treatment. Blood glucose was measured at 0 (basal), 15, 30, 45, 60 and 120 minutes from the tail vein using a glucometer (Accu-check, Roche Diagnostic, Barcelona, Spain).

ITT was performed by injecting 0.5 U/kg of insulin intraperitoneally (Humulin, France) after 10-12 hours of fasting. Blood drops were collected from the tail vein, and glucose was measured with a glucometer at 0 (basal), 15, 30, 45, 60 and 120 minutes. The glucose area under the curve (AUC) was calculated using the basal point (0) as the basal glucose level for each mouse.

### Food intake and open field (OFT) tests

Five weeks after treatment, food intake test was performed by weighing mice and food pellets for 5 days. Daily kcal consumption was calculated based on the high-fat diet. OFT was performed to measure the locomotor activity of obese mice treated with miR-21 mimic and control mimic. Mice were moved to the experimental room and kept there for 30 minutes before starting the test. Then, each mouse was placed in the middle of the open field arena for 10 minutes, and variables such as the time and distance walked in the center, entries into the center and overall distance traveled were measured using by video tracking SMART software.

### Positron emission tomography (PET)

18F-Fluorodeoxyglucose PET (18F-FDG PET) imaging was performed at the Unidad de Imagen Molecular, CIMES, Spain (Prieto *et al*, 2011). Animals (n=6; 34.33 ± 3.2 g) fasted overnight, were anesthetized by inhalation of a mixture of isoflurane/oxygen (5% for induction and 2% for maintenance) and placed prone on a PET scanner bed 1 hour after intraperitoneal radiotracer administration (10.36 ± 2.22 MBq) to perform a static acquisition of 30 minutes. Images were subsequently reconstructed using an iterative 3-D row action maximum likelihood algorithm (3-D RAMLA). Corrections for dead time decay and random coincidences were applied. Images were reconstructed on a 128 x 128 x 120 matrix, where the voxel size equals 1 x 1 x 1 mm. PET images were normalized using the whole brain average uptake with PMOD software (3.3 PMOD Technologies Ltd., Zurich, Switzerland). The inguinal AT region of interest (ROI) was manually drawn.

Animals were kept at 25°C to maintain BAT activity as low as possible and to ensure that differences in thermogenic activity were only due to treatment. The blood glucose concentration (88.83 ± 6.88 mg/dl) was determined before tracer injection using a glucose level meter with test strips (Accu-Check Aviva Nano, Roche, Mannheim, Germany).

### Histochemistry and immunofluorescence

AT samples were fixed in 4% formaldehyde for 48 hours and then placed and processed in paraffin Spin Tissue Processor STP 120 to allow the infiltration of paraffin into the tissue. Infiltrated samples were then embedded in paraffin using the paraffin embedding center (EG1150H, Leica, Nussloch, Germany). The tissue was cut into sections of up to 5 μm thick. After sample deparaffinization, nuclear staining was carried out with Harris hematoxylin, followed by cytoplasmic staining with a mixture of eosin and 0.2% glacial acetic acid. The slides were dehydrated through a series of increasing grades of ethanol solutions. All sections were photographed using an Olympus BX61 microscope (Olympus, Tokyo, Japan).

Immunofluorescence was performed by incubating the samples in PBS plus 0.3% Triton X-100, 10% donkey serum (DS) and 10% sheep serum (SS) for 1 hour. Then, the sections were incubated in a mixture of primary antibodies including rabbit anti-TMEM26 (1:50, NBP2-27334, Novus Biologicals Europe, Abingdon, United Kingdom) and goat anti-UCP1 (1:75, SAB2501082, Merck KGaA, Darmstadt, German) overnight at 4°C. The slides were incubated at room temperature for 2 hours with the fluorescent-labeled secondary anti-rabbit IgG−FITC antibodies (1:50, F7512, Merck KGaA, Darmstadt, Germany) suspended in PBS + 5% DS and then incubated at room temperature for 2 hours in anti-goat IgG H&L TRITC (1:50, ab6522, Abcam, Oxford, UK) suspended in PBS + 5% SS. Finally, Fluoroshield™ DAPI medium was added. All sections were photographed using an Olympus BX61 microscope. Fluorescence photomicrographs were captured with a digital camera (DP70, Olympus, Tokyo, Japan) and software DP Controller (1.2.1.108, Olympus, Tokyo, Japan).

### Transmission electron microscopy (TEM)

Small pieces of tissue were fixed with 2.5% glutaraldehyde solution for 75 hours. Sections were washed in phosphate buffer, pH7.4, and placed in an Automatic Sample Processor (EM. TP, Leica, Nussloch, Germany) for 25 hours and 55 minutes. The processor performs washes in PB, followed by application of 2% osmium in PB, washed in distilled water and acetone in increasing gradation from 25% to 100%. The processing continues with the inclusion in Epon resin in different percentages mixed with acetone, up to 100% pure resin. The re-cutting and semithin cuts were carried out using a glass blade in a standard range of 300 nm. To determine the areas for ultrathin cuts, the sections were placed on standard glass slides and stained with toluidine blue (89640, Merck KGaA, Darmstadt, Germany). Finally, the sections were placed on 300 mesh copper grids. The study of cell organelles was performed under a transmission microscope with a voltage of 80 kilovolts (Libra-120, ZEISS Oberkochen, Alemania), and the images were obtained using ITem software (Olympus, Olympus, Tokyo, Japan).

### miRNA extraction and real-time quantitative PCR (qPCR)

miRNA was isolated from AT samples and 3T3-L1 cells using the miRvanaTM miRNA isolation kit (Ambion, AM1561, Spain) according to the manufacturer’s protocol. For plasma samples, automated extraction was carried out using Maxwell 16 Promega and the Maxwell (R) 16 miRNA Tissue kit (Promega, AS 1470, Spain), according to the manufacturer’s recommendations. The miRNA concentration was measured using a Nanodrop ND-2000 (Thermofisher Scientific, USA) or Quantifluor RNA kit (E3310, Promega, Spain) and Quantus Fluorometer (Promega, Spain). Five nanograms of miRNA were converted to cDNA by specific reverse transcription that was carried out using specific primers for each miRNA (looped RT primer TaqMan small RNA assays), namely, hsa-miR-21 (000397), control miRNA assay snoRNA-142 (001231) and control miRNA assay RNU48 (001006), and the TaqMan MicroRNA Reverse Transcription kit (Thermofisher Scientific, USA). RT qPCR reactions were carried out using specific TaqMan probes and an Agilent Mx3005P QPCR system. During amplification, the Ct value was determined, and specific signals were normalized with stable expression (BestKeeper) of the reference gene snoRNA-142 (mouse) or RNU48 (human) using the formula 2^-Δct^.

### mRNA extraction and real-time qPCR

Total RNA was extracted from ATs with the Qiasol RNeasy Lipid Tissue mini kit (Qiagen, Valencia, CA) or with RNA-Stat 60 Reagent (Ams Biotechnology, Abingdon, UK) for 3T3-L1 cells, according to the manufacturer’s recommendations. Then, 4, 2 or 1 μg of total RNA was converted to cDNA using reverse transcriptase (Transcriptor Reverse Transcriptase 20 U/μL, 03531287001, Roche). Next, RT qPCR amplification was performed using specific TaqMan Gene Expression Assays, 10 ng of cDNA and Brilliant III Ultra-Fast QPCR master mix. Specific signals were normalized by constitutively expressed TBP (Mm 00446973, Thermofisher scientific) in AT using the formula 2^-Δct^ or β-actin (mouse ACTB, 452341E, Thermofisher scientific) in 3T3-L1 cells using the formula 2^-ΔΔct^. The references of TaqMan probes evaluated in this study are Vegf-A (Mm 00437306-m1), Vegf-b Mm 00442102_m1, Ucp1 (Mm 01244861-m1), Tmem26 (Mm 01173641-m1), Pgc-1α (Mm 01208835), Prdm16 (Mm 00712556-m1), Cidea (Mm 00432554-m1), Ppar-γ (Mm 00440940-m1), P53 (Mm01731290-g1), Tgfβ1 (Mm01178820-m1), Fgf21 (Mm 00840165-g1), Hoxc9 (Mm 00433972-m1), Sirt1 (Mm01168521-m1), Mmp-9 Mm 00442991_m1, Timp-2 Mm 00441825_m1,Timp-3 Mm 00441826-m1, Cebpα Mm 00514283-s1, Bcl-2 Mm 00477631-m1, Bid Mm 00432073_m1 and Casp3 Mm 01195085-m1

### DNA Extraction and qPCR

Total DNA was extracted from ingSAT using DNeasy Blood and Tissue Kit (Qiagen, Valencia, CA) according to the manufacturer’s recommendations. The DNA concentration was measured using a Nanodrop ND-2000 (Thermofisher Scientific, USA). Relative mtDNA content of ingSAT was analyzed by qPCR. Multiplex qPCR amplification was performed using specific probes, Brilliant III Ultra-Fast QPCR master mix, 160 ng of DNA and reference dye. The GADPH gene was used as a nuclear gene (nDNA, Endogenous Control, VIC™/MGB probe, primer limited, Thermofisher cat 4352339E) and *Cox1* as mitochondrial gene (mtDNA, Mm04225243_g1). The relative mtDNA content was calculated the formula: mtDNA= 2-Δct where ΔCt= Ct *Cox1*- Ct *Gadph*.

### Prediction of putative miRNA target genes

The miRTarBase 4.0 website (https://mirtarbase.mbc.nctu.edu.tw/), TarBase v8 (http://carolina.imis.athena-innovation.gr/diana_tools/web/index.php?r=tarbasev8 index) and the miRWalk 2.0 (https://www.umm.uni-heidelberg.de/apps/zmf/mirwalk/index.html) databases were used for prediction of validated and nonvalidated target genes (TGs) of miR-21. The Protein Analysis through Evolutionary Relationship (PANTHER) Classification System (https://www.pantherdb.org/) and the GeneCodis3 (Gene annotations co-occurrence discovery) (http://genecodis.cnb.csic.es./analysis) were applied to annotate the biological processes of the predicted targets.

### Interactions among miRNAs, biological processes, and miR-21 target genes

The interactions among miR-21, biologic processes, and the TGs were visualized with Cytoscape version 3.2.1 software (https://www.cytoscape.org/).

### Statistical analyses

Statistical analyses and graphics were carried out using GraphPad Prism 5.00.288 and IBM SPSS Statistics 22. All data are expressed as the mean ± SEM. Comparisons between multiple groups were performed by one-way ANOVA with Bonferroni’s post hoc test, and comparisons between two groups were performed by Student’s t-test. GTT, ITT and body weight were analyzed by repeated two-way ANOVA measurements. p□0.05 was considered statistically significant. In the PET study, significant differences between groups were determined using a 2-tailed 2-sample unequal variance Student’s t-test in Microsoft Excel. A p<0.05 was considered to indicate statistical significance.

## Data Availability

Yes data are available

## Acknowledgments

The authors wish to thank all the subjects for their collaboration. CIBER Fisiopatología de la Obesidad y Nutrición (CIBEROBN) is part of the ‘Instituto de Salud del Carlos III’ (ISCIII). Project. This work was supported in part by grants from the Instituto de Salud Carlos III (ISCIII)/FEDER-UE (PI18/00785), Consejería de Salud, Junta de Andalucía (PI-0092-2017), Spain and co-funded by the Fondo Europeo de Desarrollo Regional (FEDER). R.E.B. and F.J.B.S. are under a contract from the ‘Nicolas Monarde’ (C-0030-2016, RC-0005-2016) program from the Servicio Andaluz de Salud, Regional Ministry of Health of the Andalusian Government, Andalusia, Spain. M.C.P. was a recipient of a post-doctoral grant Juan de la Cierva Formación (FJCI-2017-32194) from the Ministerio de Ciencia, Innovación y Universidades (Spain).

## Author Contributions

R.E.B. and F.J. T. designed the research. R.E.B. performed experiments, analyzed the data and wrote the paper. S.L. and A.M.G. performed *in vivo, in vitro, ex vivo* and molecular biology experiments and bioinformatics analysis. W.O.O. carried out *in vitro* culture of preadipocytes. R. M.G.P. and S. L. carried out histologic and microscopic analysis. M. F.C. carried out PET analysis. S.Y. R.Z. V. E., and F.J. B.S. carried out animal models of obesity generation. C.L.G performed mtDNA analyzes. H. Z. contributed to manuscript writing and editing. M.C.P. and N. H. contributed to statistical, data analysis and interpretation. G.O.F. and J.S. contributed to human subjects’ recruitment and clinical analysis.

## Conflict of interest

The authors have no conflicts of interests to declare.

## Notes

### Competing Interest Statement

The authors have declared no competing interest.

### Funding Statement

The authors wish to thank all the subjects for their collaboration. CIBER Fisiopatologia de la Obesidad y Nutricion (CIBEROBN) is part of the Instituto de Salud del Carlos III (ISCIII). Project. This work was supported in part by grants from the Instituto de Salud Carlos III (ISCIII)/FEDER-UE (PI18/00785), Consejeria de Salud, Junta de Andalucia (PI-0092-2017), Spain and co-funded by the Fondo Europeo de Desarrollo Regional (FEDER). R.E.B. and F.J.B.S. are under a contract from the Nicolas Monarde (C-0030-2016, RC-0005-2016) program from the Servicio Andaluz de Salud, Regional Ministry of Health of the Andalusian Government, Andalusia, Spain. M.C.P. was a recipient of a post-doctoral grant Juan de la Cierva Formacion (FJCI-2017-32194) from the Ministerio de Ciencia, Innovacion y Universidades (Spain).

### Author Declarations

The experimental protocol was approved by the Ethics and Research Committee of the Virgen de la Victoria Clinical University Hospital (Malaga, Spain). The European Union recommendations (2010/63/EU) on animal experimentation were followed. All procedures were approved by the ethics committee of the University of Malaga (authorization no. 2012 0061A).

