## supplemental tables and figures for "miR-21 blocks obesity in mice: a potential therapy for humans"

**Running title: miR-21-induced browning activation**

Said Lhamyani ^1^**^#^**, Adriana-Mariel Gentile ^1^**^#^**, Rosa M. Giráldez-Pérez^2^, Mónica Feijóo-Cuaresma ^3^, Silvana Yanina Romero-Zerbo ^1,4^, Mercedes Clemente-Postigo ^5^, Hatem Zayed ^6^, Wilfredo Oliva Olivera^6^, Francisco Javier Bermúdez-Silva ^1,4^, Julián Salas ^8^, Carlos López Gómez^9^, Nabil Hajji ^10^, Gabriel Olveira Fuster ^1,4^, Francisco J. Tinahones ^6^, Rajaa El Bekay ^1,11^.

^1^Unidad de Gestión Clínica de Endocrinología y Nutrición, Instituto de Investigación Biomédica de Málaga (IBIMA), Hospital Regional Universitario, Universidad de Málaga, Málaga, Spain.

^2^Biología Celular, Fisiología e Inmunología. Universidad de Córdoba. Córdoba, Spain

^3^Unidad de Imagen Molecular (UIM, www.uimcimes.es). Centro de Investigaciones Médico-Sanitarias (CIMES) de la Universidad de Málaga.

^4^CIBER de Diabetes y Enfermedades Metabólicas Asociadas (CIBERDEM), Málaga, Spain.

^5^Department of Cell Biology, Physiology, and Immunology; Maimónides Biomedical Research Institute of Córdoba (IMIBIC)/University of Córdoba/Reina Sofia University Hospital; Córdoba, Spain.

^6^Unidad de Gestión Clínica de Endocrinología y Nutrición, Instituto de Investigación Biomédica de Malaga (IBIMA), Hospital Universitario Virgen de la Victoria, CIBER Fisiopatología de la Obesidad y Nutrición (CIBERobn), Málaga, Spain.

^6^Department of Biomedical Sciences, QU Health, College of Health Sciences, Qatar University, Doha, Qatar

^8^Cardiovascular Surgery Department, Carlos Haya University Hospital, Malaga, Spain

^9^Unidad de Gestión Clínica de Aparato Digestivo, Instituto de Investigación Biomédica de Málaga (IBIMA) Hospital Universitario Virgen de la Victoria. Málaga.

^10^ John Fulcher Neuro-Oncology Laboratory, Division of Brain Sciences, Imperial College London, London, UK.

^11^CIBER-The Spanish Biomedical Research Centre in Physiopathology of Obesity and Nutrition, Institute of Health Carlos III, Malaga, Spain.

**^#^Contributed equally**

**Corresponding Authors:**

El Bekay R:

Tinahones FJ:

| **Table S1:** **DAVID bioinformatic analysis. Biological Functions of miR-21´s targets** | | |
| --- | --- | --- |
| **Biological Functions** | **p Value** | **Target genes** |
| Brown fat cell differentiation | 0.0821 | *CEBPA, ADRB2, CEBPB, PTGS2, PPARGC1A* |
| Regulation of fat cell differentiation | 0.0409 | *AKT1, ALMS1, LPIN1, FNDC3B, SOD2* |
| Angiogenesis | 0.0499 | *WARS, VEGFC, COL4A1, AGGF1, VEGFA, APOLD1, RHOB, ARHGAP24, MMP2, EPHA2* |
| Regulation of angiogenesis | 0.0380 | *WARS, HIF1A, AGGF1, ERBB2, NF1, TEK, IL1B, RHOB, THBS1, VASH2, DDAH1* |
| VEGF signaling pathway | 0.00249 | *AKT1, NRAS, PTK2, KRAS, PTGS2, RAC1, MAPK3, VEGFA, PIK3CA, NOS3, BAD, PIK3R1, AKT2* |
| Apoptosis | 0.0115 | *RTN4, E2F1, MEF2C, E2F2, MEF2A, CADM1, ZMAT3, FASLG, TP63, NFKB1, FOXO3, DAXX, PDCD4, PTEN, CTNNB1, SMNDC1, AKT1, BAG4, TRIAP1, TNFRSF11B, PAK2, SLK, TIAM1, CASP8, IL1B, RHOB, NGFRAP1, FAS, MYC, TOP2A, NDUFS1, RPS27A, CCAR1, BCL10, TP53, HRK, TOPORS, ARHGEF12, ECT2, PPM1F, TNFRSF10B, CDK11A, BNIP2, HIPK3, HIPK2, MDM4, TNFAIP3, EIF2AK3, BID, APH1A, CLU, RFFL, RRAGC, PEA15, FIS1, TSC22D3, MOAP1, DOCK1, CSE1L, BCL2, MAP3K1, SOS2, TRAF7, THBS1, AXIN1, HIP1, DNM1L, MSH2, TP53BP2, RYBP, ALMS1, BIRC5, SOD1, BRCA1, FXR1, SOD2, VDAC1, NCSTN, PHF17, NRAS, GSPT1, MAP1S, APAF1, PDCD6, F2R* |
| Insulin receptor signaling pathway | 0.00297 | *PHIP, IGF1R, EIF4EBP2, GAB1, AP3S1, RHOQ, FOXO1, APPL1, PIK3R1, AKT2* |
| Insulin signaling pathway | 0.0637 | *PRKAB2, SOCS1, HK2, MKNK2, FOXO1, RHOQ, SOCS4, PPARGC1A, AKT1, MAPK1, NRAS, CRKL, TSC1, INPP5K, SOS2, PRKAA2, RAPGEF1, PIK3R1, AKT2* |
| MiRTarBase7.0, MiRWalk 2.0, and Tarbase 8.0 used for the prediction of validated target genes of miR-21. A total of 1505 of validated target genes for miR-21 were introduced in the DAVID database to annotate the biological functions in which they are involved. | | |

| **Table S2: Panther bioinformatic analysis. Biological processes of miR-21´s target** | | | |
| --- | --- | --- | --- |
| **Biological Process** | **Number of genes** | **Percent of gene hit against total genes** | **Percent of gene hit against total process hits** |
| Cellular process | 511 | 34.0% | 29.2% |
| Metabolic process | 413 | 27.4% | 23.6% |
| Biological regulation | 307 | 20.4% | 17.5% |
| Localization | 183 | 12.2% | 10.4% |
| Response to stimulus | 102 | 6.8% | 5.8% |
| Multicellular organismal process | 92 | 6.1% | 5.2% |
| Developmental process | 38 | 2.5% | 2.2% |
| Immune system process | 32 | 2.1% | 1.8% |
| Biological adhesion | 23 | 1.5% | 1.3% |
| Cellular component organization or biogenesis | 19 | 1.3% | 1.1% |
| Cell proliferation | 15 | 1.0% | 0.9% |
| Reproduction | 13 | 0.9% | 0.7% |
| Rhythmic process | 3 | 0.2% | 0.2% |
| Biological phase | 1 | 0.1% | 0.1% |
| Pigmentation | 1 | 0.1% | 0.1% |
| MiRTarBase7.0, MiRWalk 2.0, and Tarbase 8.0 used for the prediction of validated target genes of miR-21. A total of 1505 of validated target genes for miR-21 were introduced in the PANTHER database to annotate the biological processes in which they are involved. | | | |

**Table S3**: **Binding sites of miR-21 within the objective.**

| GENE | ID | 3' UTR length | miRNA | Position | seed match | Context ++ score | Context ++ score Percentil | Pct |
| --- | --- | --- | --- | --- | --- | --- | --- | --- |
| UCP1 | ENST00000262999.3 | 464 | hsa-miR-21-5p | 245-252 | 8mer | -0.34 | 97 | <0.1 |
| TP53 | ENST00000420246.2 | 1496 | hsa-miR-21-3p | 59-66 | 8mer | -0.37 | 98 | N/A |
| HOXC9 | ENST00000593756.1 | 114 | hsa-miR-21-3p | 195-201 | 7mer-m8 | -0.28 | 96 | N/A |
| TP53 | ENST00000420246.2 | 1496 | hsa-miR-21-3p | 460-466 | 7mer-m8 | -0.08 | 67 | N/A |
| PPPGC1A | ENST00000264867.2 | 3801 | hsa-miR-21-3p | 2055-2061 | 7mer-m8 | -0.02 | 22 | N/A |

**Table S4.** **Anthropometric and biochemical characteristics of the study groups.** The clinical-pathological and biochemical variables of healthy normoweight subjects (NW), morbidly obese with low insulin resistance patients (LIR-MO), and morbidly obese patients with high insulin resistance (HIR-MO).

|  | **NW**  **(n=7)** | **LIR-MO**  **(n=9)** | **HIR-MO**  **(n=9)** |
| --- | --- | --- | --- |
| **Age (years)** | 42.29 ± 5.08a | 43.44 ± 3.87a | 39.00 ± 2.78a |
| **BMI (kg/m^2^)** | 26.15 ± 0.88a | 55.68 ± 2.14b | 57.52 ± 1.93b |
| **Triglycerides**  **(mg/dL)** | 88.71 ± 12.28a | 117.33 ±17.88a | 139.08 ± 21.41a |
| **HOMA index** | 2.91 ± 0.35a | 2.62 ± 0.25a | 12.26 ± 1.19b |
| **Total Cholesterol**  **(mg/dL)** | 192.29 ± 5.95a | 198.29 ± 10.74a | 185.89 ± 16.03a |
| **Cholesterol LDL** **(mg/dL)** | 111.54 ± 9.07a | 124.22 ±12.90a | 137.66 ±33.86a |
| **Cholesterol HDL**  **(mg/dL)** | 64.00 ± 4.80a | 50.44 ± 4.10b | 43.67 ± 3.55b |
| **Waist**  **Circumference (cm)** | 94.86 ± 2.84^a^ | 138.67± 6.15^b^ | 145.67 ± 8.28^b^ |
| **Glucose mg/dL** | 98.43±2.95^a^ | 96.78±2.74^a^ | 114.56±56^b^ |
| **Insulin**  **U/ml** | 11.85±1.19^a^ | 10.94±0.95^a^ | 43.66±3.14^b^ |

Donors (n = 25) were selected according to BMI and glycemic state. Data are expressed as the mean ± SEM. Comparison among groups was performed by one-way ANOVA with Bonferroni´s PostHoc test. Different superscript letters represent statistically significant differences between groups (p<0.05). BMI: body mass index, HOMA index: homeostasis model assessment index;

**Figure S1. Body weight gain and glucose tolerance test of a control diet and 45% HFD mice previously to the start of the miR-21 *in vivo* treatment**.

Twice a week, body weight was monitored in awake mice. After 14 weeks of diet, glucose tolerance was assessed by intraperitoneal glucose tolerance test (GTT) for the two groups (control diet and 45% HFD). AUC was calculated using as the basal point (0) the weight before miR-21 mimic treatment or basal glucose level before the GTT for each mouse. The GTT was carried out by injecting 2 g/kg of D-glucose or 0.5 U/kg of insulin after 10-12h of fasting. Blood samples were collected from the tail vein at 0 (basal), 15, 30, 45, 60, and 120 min, and glucose was measured at the set times with a glucometer. Data are expressed as mean ±SEM. *p˂0.05, **p<0.01 versus control diet. &p˂0.05 versus basal point (0) according to repeated measures ANOVA and student´s t-test, respectively.

**Figure S2**. **miR-21 levels in ingWAT, BAT, VAT, and plasma from miR-21 mimic treated mice.** miR-21 levels were measured and analyzed in ingWAT, BAT, VAT, and plasma from obese mice treated with 0.5 µg of mimic miR-21 or control mimic (n=7) by real-time qPCR using SnoRNA142 (adipose tissue) and U6 (plasma) as reference genes (2^-Δct^). Data are expressed as mean ±SEM. *p˂0.05, **p˂0.01 versus control mimic according to student´s t-test.

**Figure S3. Generation of diet-induced non-diabetic and diabetic obese mice and study design for miR-21 expression analyses.**

**(A)** C57BL/6J mice (11-weeks old at arrival) were separated into three groups: (1) mice fed with control diet (n=9) (containing 10% of kcal from saturated fat) (control) from the day 0 (start of the experiment) to 14 weeks (end of the experiment); (2) non-diabetic obese mice (45% HFD-ob; n=8) fed the control diet for 6 weeks and then, fed a diet containing 45% of kcal from saturated fat (45% HFD), for further 8 weeks; and (3) diabetic obese mice (45% HFD-diabetic; n=8) fed for 14 weeks a 45% HFD. (**B)** Body weight was monitored twice a week for the duration of the experiment. (**C**) Glucose (GTT) and insulin (ITT) tolerance tests were carried out during the last week of diet by injecting 2 g/kg of D-glucose or 0.5 U/kg of insulin after 10-12 hours of fasting. Blood samples were collected from the tail vein at 0 (basal), 15, 30, 45, 60, and 120 minutes, and glucose was measured at the set times with a glucometer. Data are expressed as the mean ± SEM. *p˂0.05 versus control diet according to repeated measures ANOVA test.

**Figure S1**

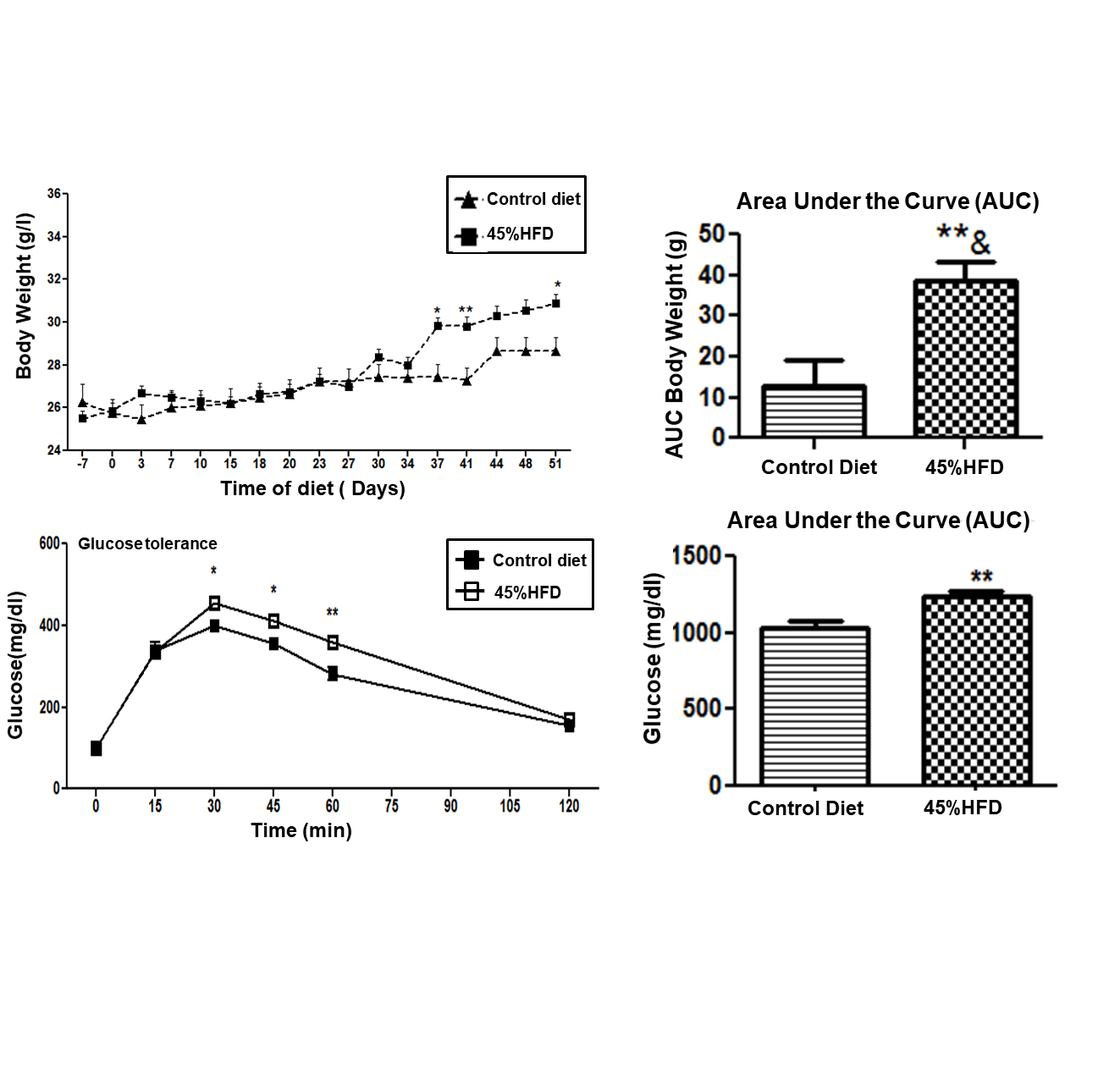

**Figure S2**

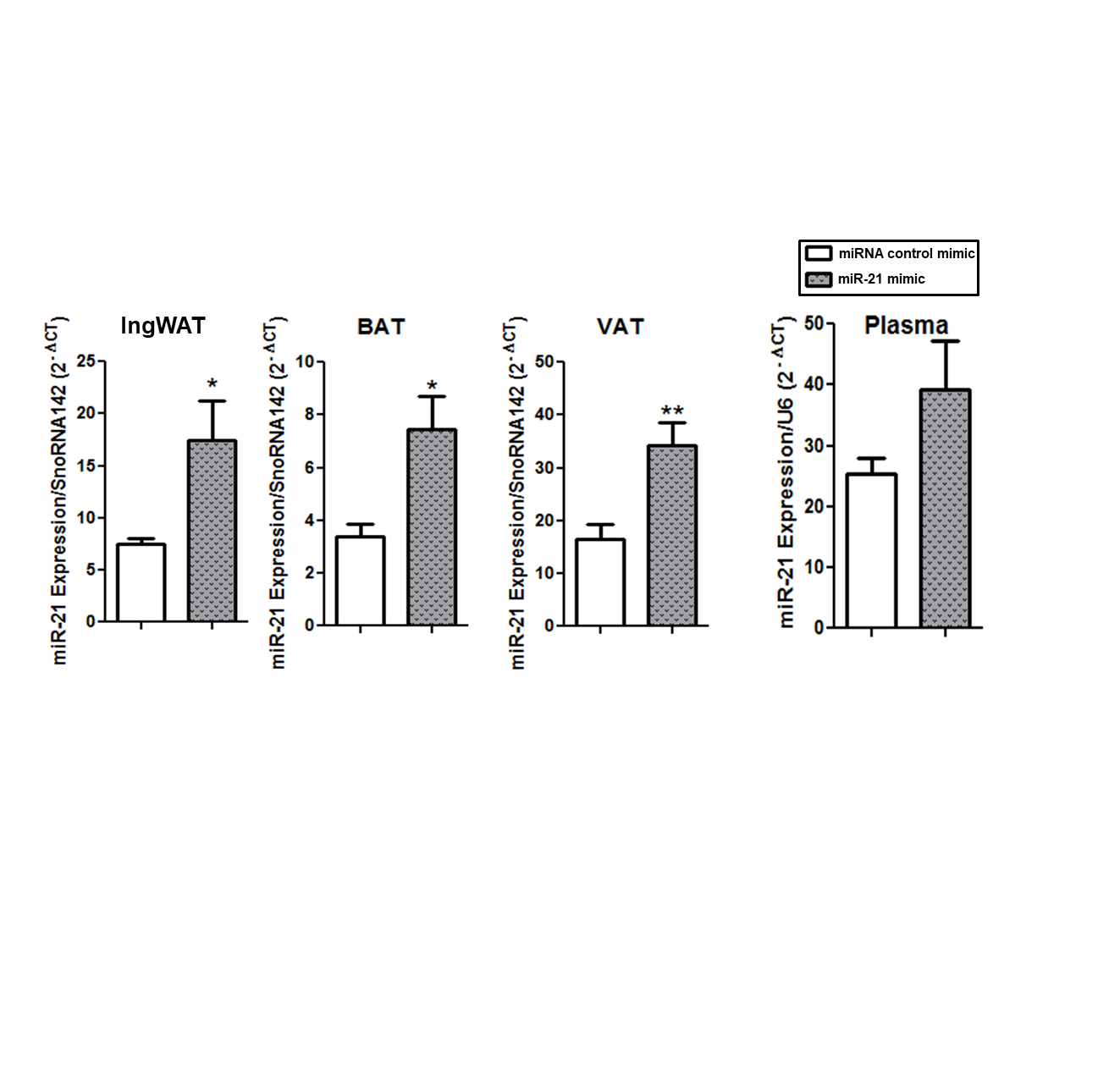

**Figure S3.**

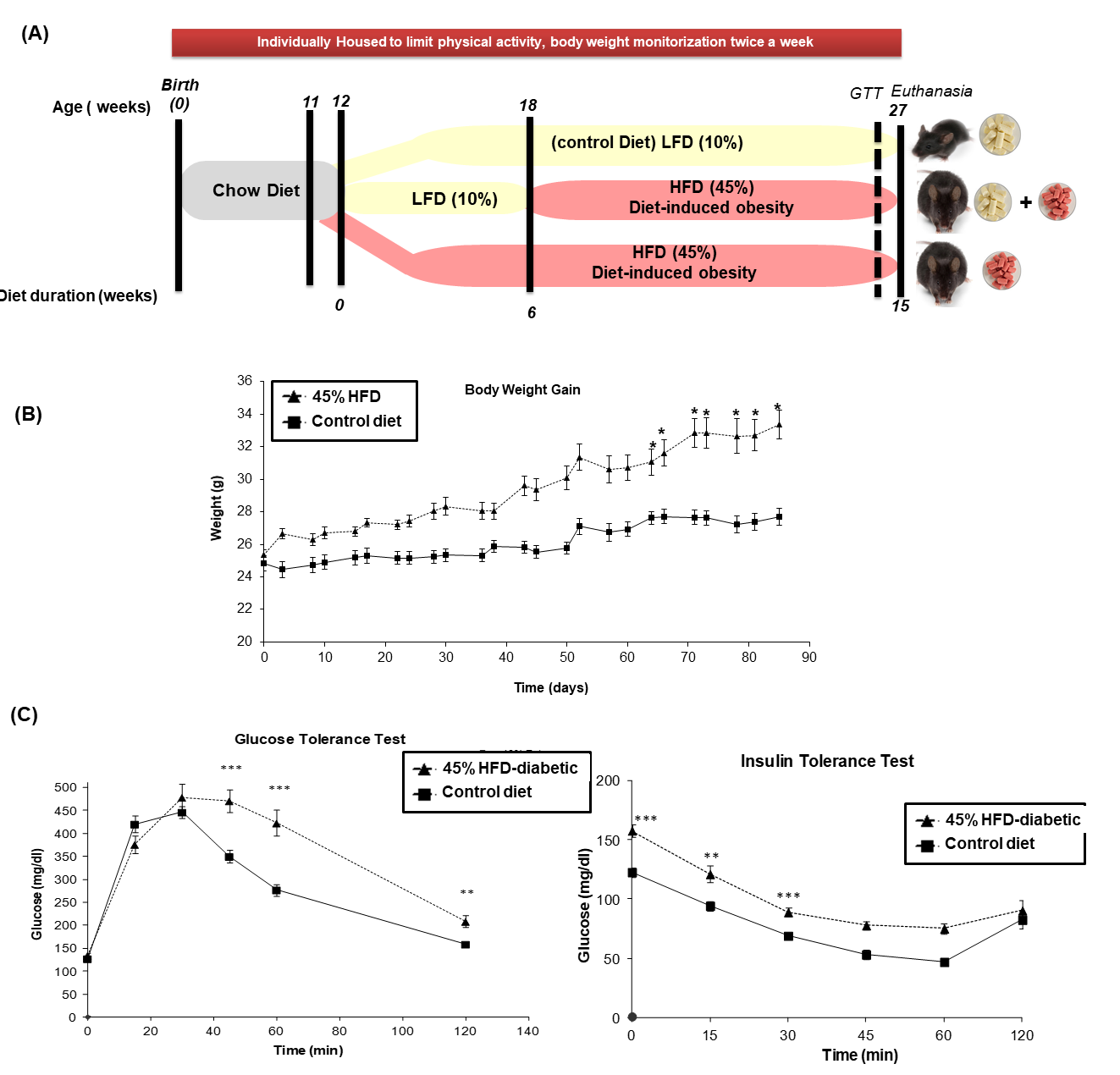
